## Supplementary appendix for "The impact of different antimicrobial exposures on the gut microbiome in the ARMORD observational study"

#### Supplementary methods

##### Laboratory methods

DNA extraction was performed using bead beating followed by QIAGEN Fast DNA Stool Minikit using the following protocol:

*Preparation*
1) Prepare 2ml Lysing Matrix E tube (MP Biomedicals) by adding 1ml Stool Transport and Recovery buffer (Roche) and *T. thermophilus* DNA (DSMZ, typically 100ng, but amount varied between extractions). Label and weigh tubes.
2) Remove stool samples from -80˚C to thaw immediately before extraction.

*Lysis*
3) Add ~20–1000mg sample to each tube, transferring more from more liquid samples.
Weigh again to establish weight of sample.
4) Bead beat tubes twice at 6m/s for 40s with a FastPrep-24 5G instrument (MPBiomedicals) with 5 minutes interval at 4˚C

5) Incubate tubes at 95˚C for 5 minutes and centrifuge at 1,000g for 1 minute
6) Transfer 900µl supernatant to 1ml InhibitEX buffer in 2ml Eppendorf, invert x 100 and centrifuge at 17,000g for 3 minutes
7) Transfer 400µl supernatant to 30µl proteinase K in 1.5ml Eppendorf
8) Add 400µl AL lysis buffer and invert x 100
9) Incubate tubes at 70˚C for 1 minute and centrifuge at 17,000g for 30s
10) Add 400µl 100% ethanol, invert x 20 and centrifuge at 17,000g for 30s

*DNA binding & washing*
11) Transfer 1st half of supernatant to spin column, centrifuge at 17,000g for 1 minute and transfer column to new collection tube
12) Transfer 2nd half of supernatant to same spin column, centrifuge at 17,000g for 1 minute and transfer column to new collection tube
13) Add 500µl AW1 wash buffer to column, centrifuge at 17,000g for 1 minute and transfer column to new collection tube
14) Add 500µl AW2 wash buffer to column, centrifuge at 17,000g for 1 minute and transfer column to new collection tube
15) Centrifuge at 17,000g for 1 minute to dry and transfer column to 1.5ml Eppendorf
Elution
16) Add 50µl molecular water and incubate at 55˚C for 10 minutes before centrifuging at17,000g for 1 minute to elute DNA

Following extraction, DNA concentration was measured with Picogreen and Qubit fluorimeters. DNA was transferred to 96-well plates and stored at -20˚C until sequencing.

Laboratory protocols were designed to minimise the potential for DNA contamination, for example only opening samples in a cabinet, minimising aerosol generation, and never having 2 sample tubes open simultaneously. Two negative controls were included in each extraction & sequencing run. Samples and negative controls were spiked with a fixed mass of DNA from *Thermus thermophilus* (an extremophile bacterium not found in human flora) to allow quantification of extraneous DNA contamination. The median relative abundance of contaminating DNA in sequenced samples was estimated to be $6.1x10^{-5}$, which was considered to have a negligible impact on microbiome measures.

#### Bioinformatic methods
SAMtools (v1.7) was used to reformat and merge sequence data, and BBDuk (v37.90) was used to remove Illumina adapters and quality-trim reads using a phred-score cut-off of 10. Human DNA reads were identified using the Kraken2 taxonomic classifier and removed prior to analysis (v2.0.6). Quality control metrics were assessed with FastQC (v0.11.7) and MultiQC (v1.5). All samples were subsampled to a depth of 3.5 million paired reads using BBDuk, and samples with fewer reads were not used in this analysis. Taxonomic classification was also performed with MetaPhlAn2 (v2.9.20 using database v292) using a standard database containing Bacteria, Archaea and viruses from RefSeq, plus the human genome. This was used to calculate the Shannon diversity index for each sample (the sum of p*ln(p) for all species, where p is proportional abundance). Kraken2 was used to assess the abundance of specific bacterial taxa. AMR gene detection in metagenomic sequence data

was performed with the ARIBA software package (v2.11.1), using the CARD database and ontology (v3.0.2). Analysis was performed in R v4.2.3 with the ontologyIndex package (v2.4).

*Outcomes*

The following bacterial taxa of interest identified using Kraken2:

1) Enterobacteriaceae (NCBI taxonomy ID 543)
2) Enterococcus (ID 1350)
3) Bacteroidetes (ID 976)
4) Clostridia (ID 186801)
5) Actinobacteria (ID 201174)

The following AMR gene classes of interest were identified using ARIBA using the Comprehensive Antibiotic Resistance Database (CARD):

1) Beta-lactamases (TEM [CARD accession ARO:3000014], SHV [ARO:3000015], CTX-M [ARO:3000016], and OXA [ARO:3000017])
2) Tetracycline-resistant ribosomal protection proteins (ARO:0000002)
3) Aminoglycoside transferases (AACs [ARO:3000121], ANTs [ARO:3000218], and APHs [ARO:3000114])
4) Macrolide resistance genes (Erm 23S ribosomal RNA methyltransferases [ARO:3000560], and Mef efflux pumps [ARO:3000747])
5) VanA (ARO:3000010)

#### Statistical methods

A representation of the antimicrobial exposure model is shown below (**Supplementary Figure 1**). This hypothetical patient has received four courses of treatment with three different agents prior to sample collection. At the time of sample collection the patient is being treated with drug B alone. Treatment courses are defined as running from the time of the initial dose until the final dose plus one dosing interval (which ranges from 6 to 24 hours depending on the agent). The sum of the area under the curve for each agent is used to calculate the total exposure to that agent:

$$\int_0^x 2^{-x/\lambda} dx = \frac{1 - 2^{-x/\lambda}}{ln2}$$

In which λ is the microbiome disruption half-life, and x is time before sample collection. The total area under the curve is defined as 1 (i.e. exposures above are multiplied by *ln*2), so exposure to each antimicrobial is in the range 0-1. Exposures to the same antibiotic at separate times are added to provide the total exposure (so in this example exposure to antimicrobial B is the sum of two areas shaded red). Exposures to different antimicrobials are independent of one another, as are exposures to the same antimicrobial via different routes of administration.

A single value of λ was used for all analyses, chosen as the common value across all antimicrobial exposures with the lowest Akaike Information criterion across 1 to 14 days in the cross-sectional model with an outcome of Shannon diversity (this model was used to derive λ because the cross-sectional model had the greatest amount of exposure data, and Shannon diversity was calculable without imputation for all samples). The optimal value of λ was 6 days (**Supplementary Table 1**), which was used for all subsequent analyses. The exposure for samples taken at various timepoints during or after a course of antimicrobials is shown in **Supplementary Figure 2**.

#### Supplementary Figure 1 – Depiction of antimicrobial exposure model

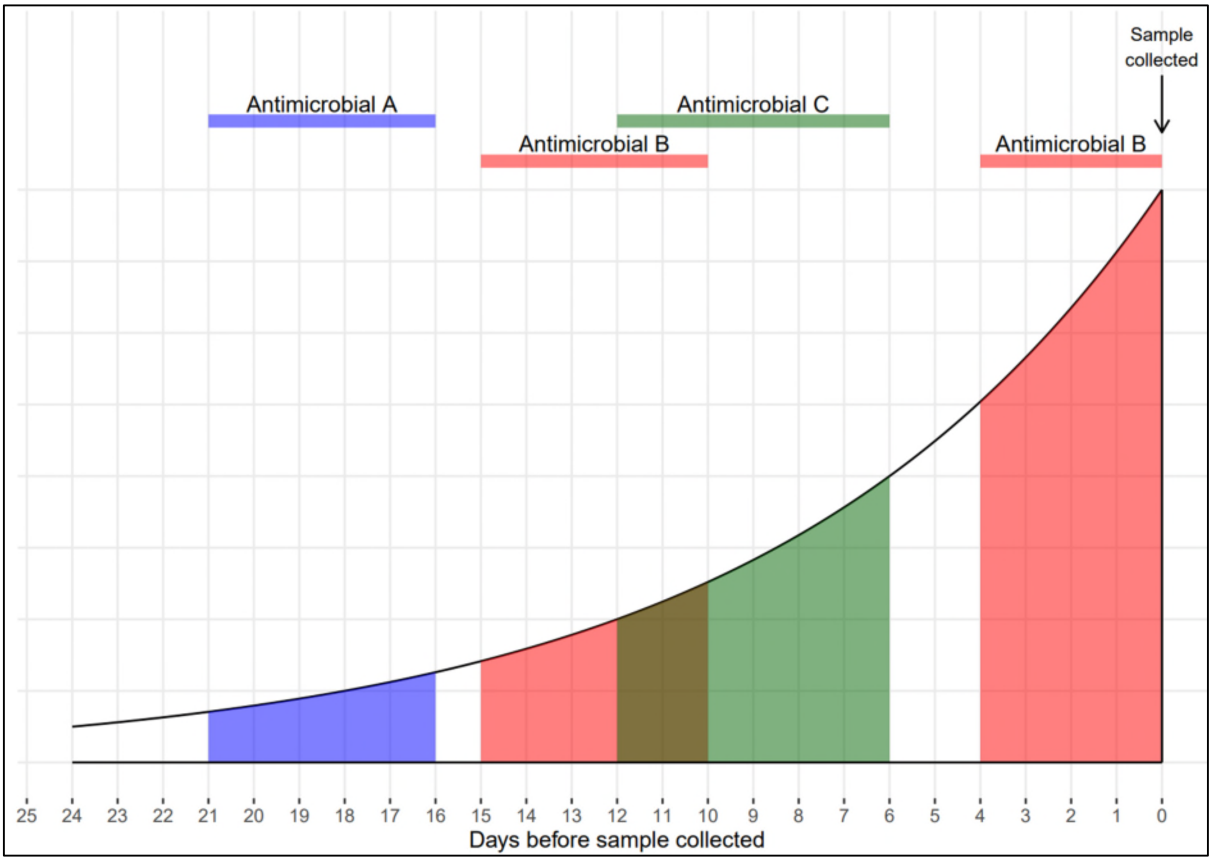

Modelled exposure for each antimicrobial is equal to the sum of shaded areas. The microbiome disruption half-life (lambda) in this example is six days.

**Supplementary Figure 2 – Modelled exposure to antimicrobial courses of varying duration.**

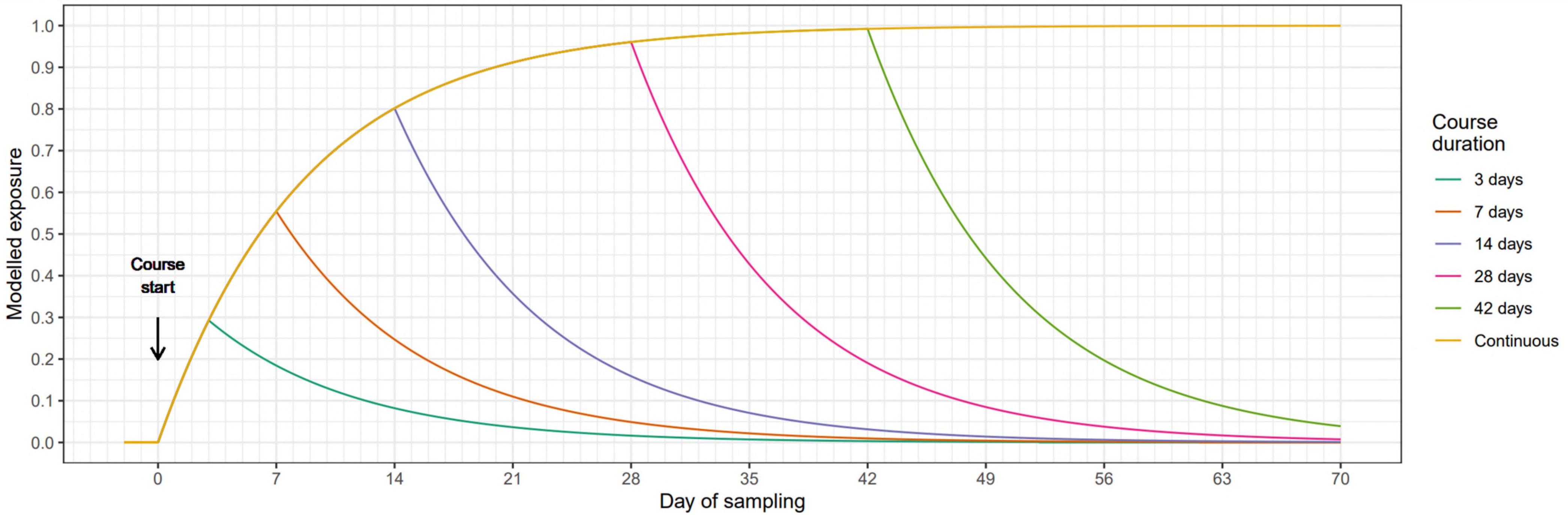

Modelled exposure to a single course of varying duration, starting at day 0. The microbiome disruption half-life (lambda) in this example is six days.

#### Supplementary Figure 3 – ARMORD study CONSORT diagram

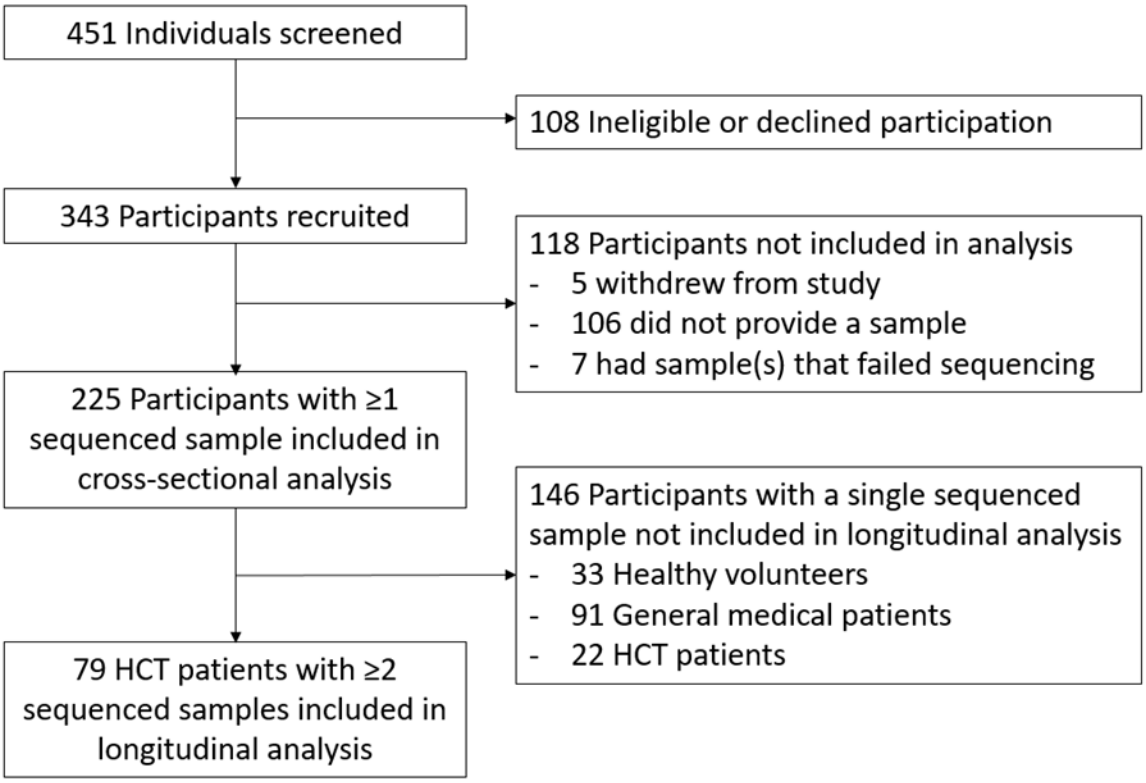

HCT = Haematopoietic stem cell transplant

**Supplementary Figure 4 - Independent effects of exposure to different antimicrobial classes on Shannon diversity in A) cross-sectional, and B) longitudinal analysis.** Figure on Multivariable estimates are in black, univariable (unadjusted) estimates in grey. Error bars represent 95% confidence intervals. Non-antimicrobial covariates are not shown, but were included in the model and can be found in supplementary data. 'Narrow' beta-lactams are penicillin, amoxicillin, flucloxacillin and first generation cephalosporins, all others are defined as 'broad'. Estimates represent the impact of prolonged use, when exposure ≈ 1 (approximately 42 days, see Supplementary Figure 2).

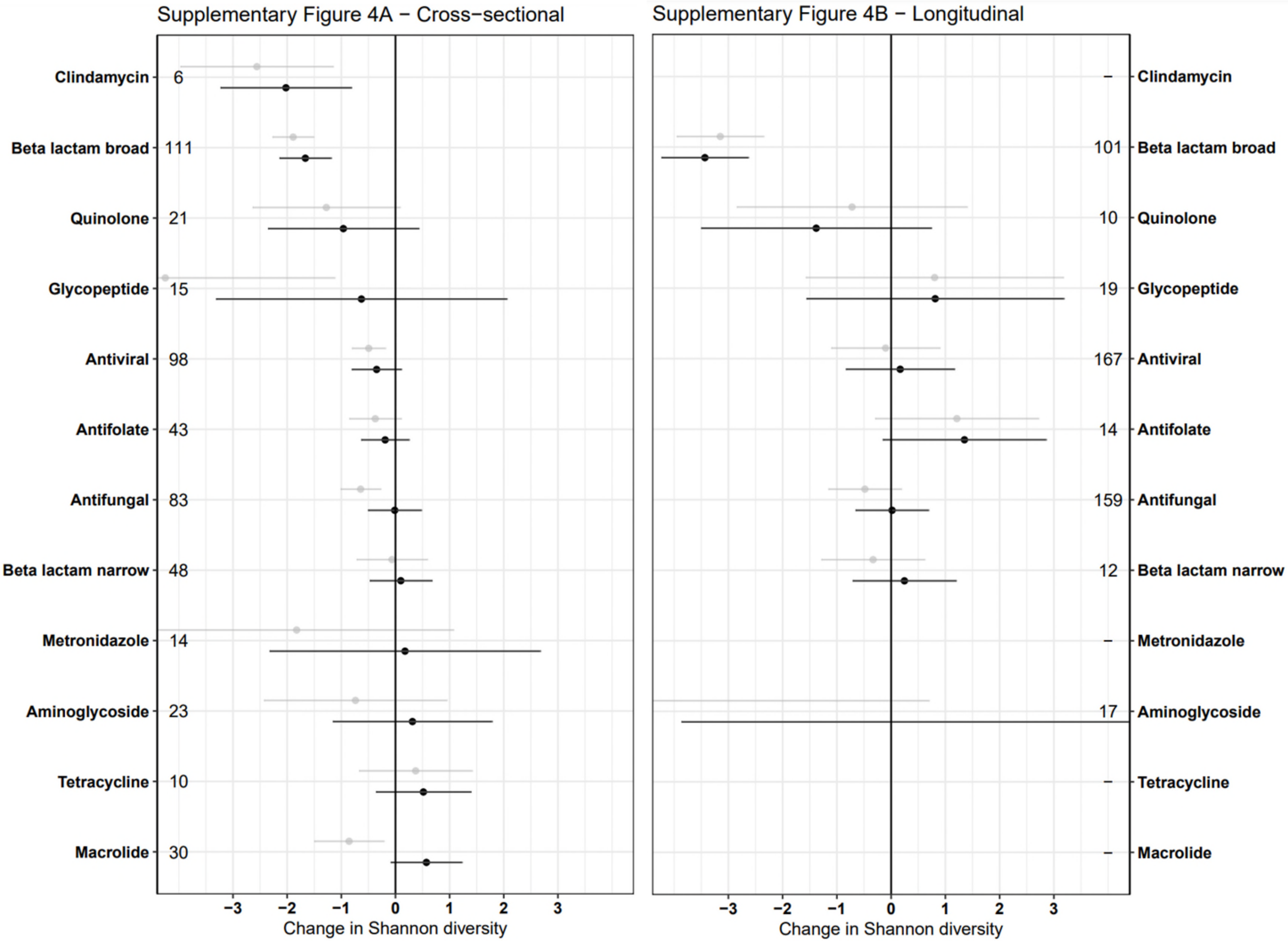

**Supplementary Figure 5 - Independent effects of exposure to different antimicrobial classes on relative abundance of selected taxa in A) cross-sectional, and B) longitudinal analysis.**
Error bars represent 95% confidence intervals. Non-antimicrobial covariates are not shown but were included in the model and can be found in supplementary data. Antimicrobial categories are the same as Supplementary Figure 4. Estimates represent the impact of prolonged use, when exposure ≈ 1 (approximately 42 days, see Supplementary Figure 2).

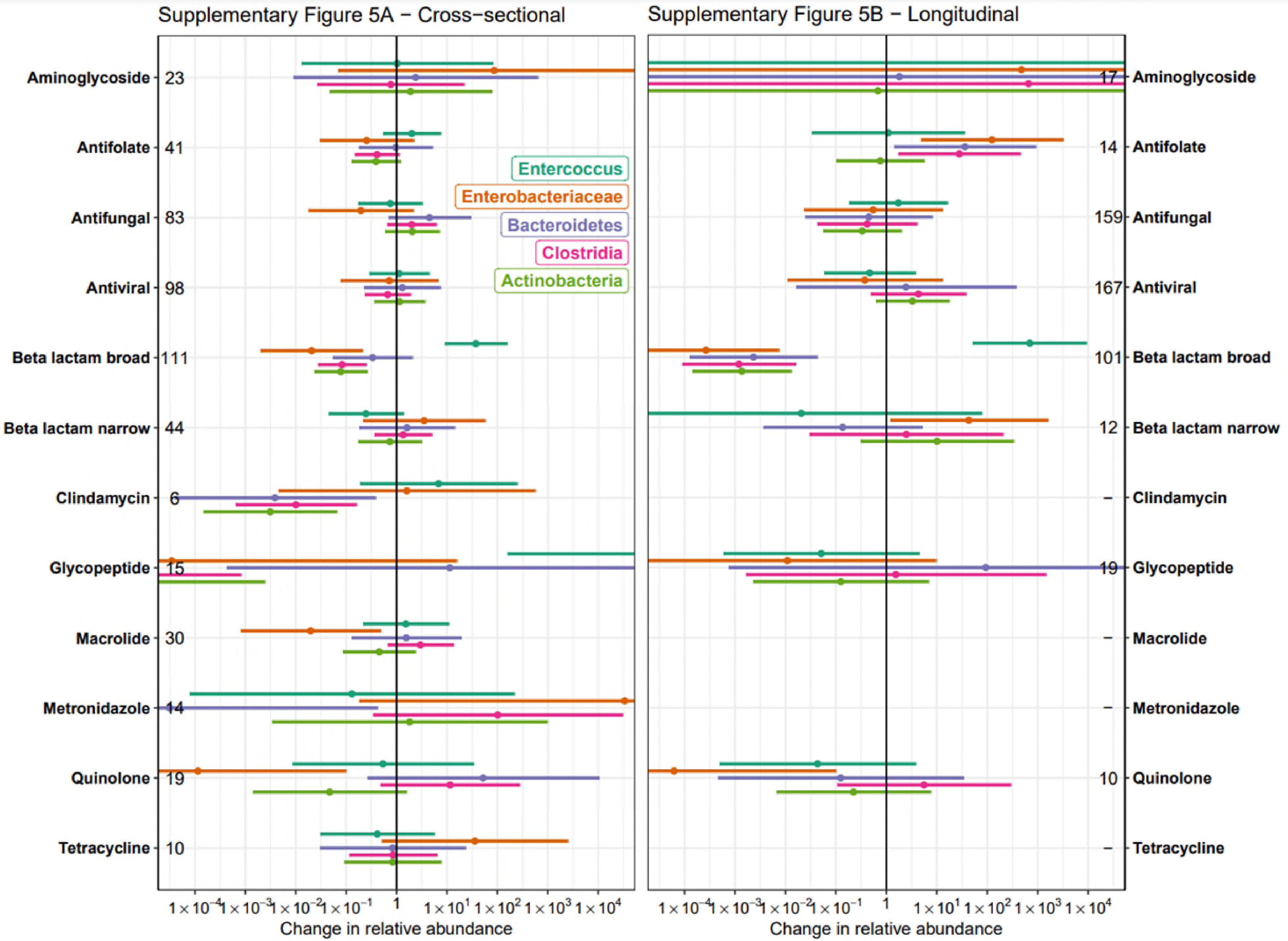

**Supplementary Figure 6 - Independent effects of exposure to different antimicrobial classes on relative abundance of selected AMR genes in A) cross-sectional, and B) longitudinal analysis.**
Error bars represent 95% confidence intervals. Non-antimicrobial covariates are not shown but were included in the model and can be found in supplementary data. Antimicrobial categories are the same as Supplementary Figure 4. Estimates represent the impact of prolonged use, when exposure ≈ 1 (approximately 42 days, see Supplementary Figure 2).

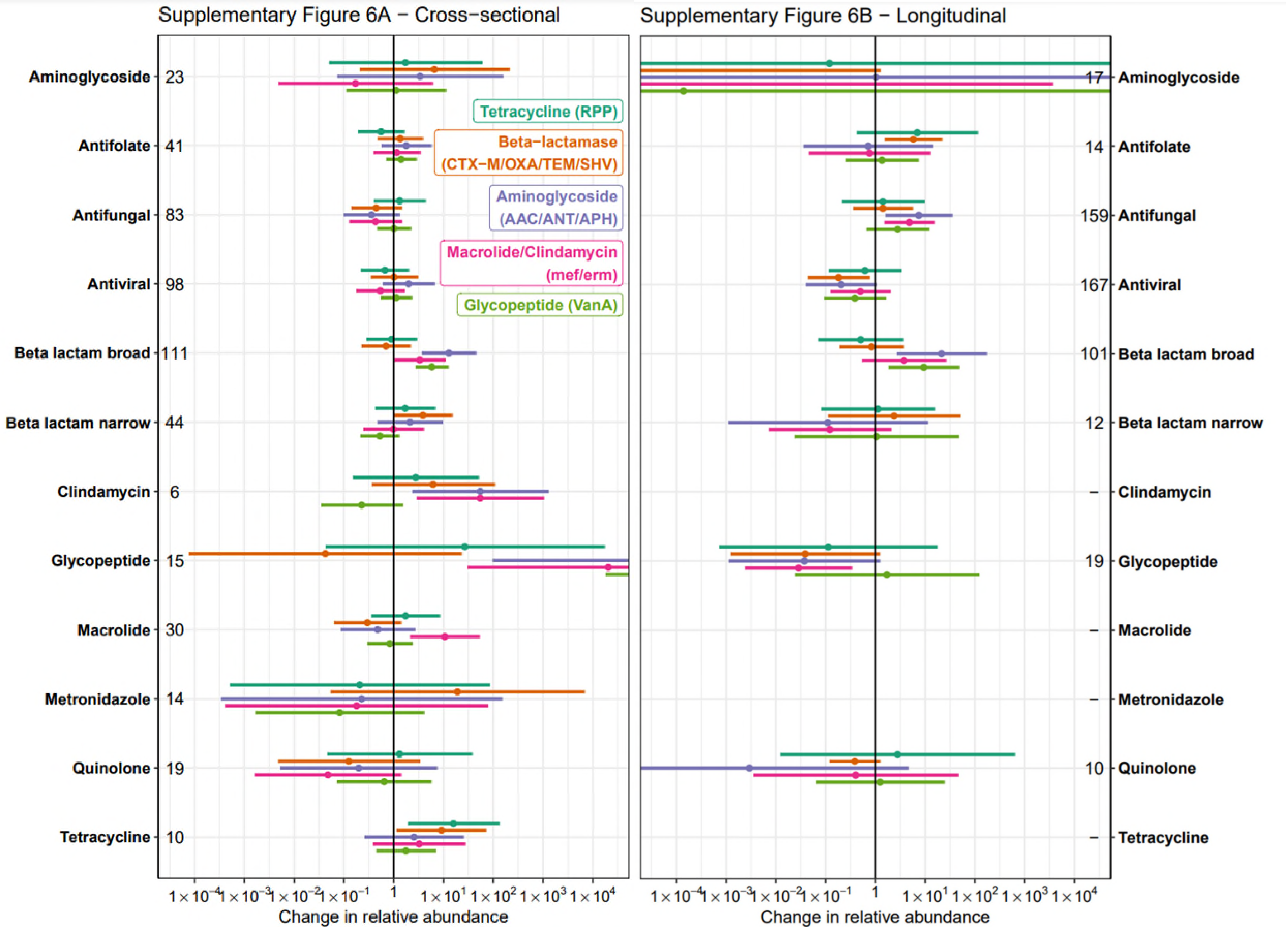

#### Supplementary Table 1 - Identification of best-fit antimicrobial exposure half-life

| Model half-life (days) | $R^2$ | Adjusted $R^2$ | Akaike Information Criterion (AIC) |
|---|---|---|---|
| 1 | 0.4078 | 0.3162 | 477.3 |
| 2 | 0.4331 | 0.3351 | 473.47 |
| 3 | 0.4583 | 0.358 | 467.24 |
| 4 | 0.4676 | 0.369 | 463.35 |
| 5 | 0.4747 | 0.3774 | 460.33 |
| **6** | **0.505** | **0.4102** | **448.95** |
| 7 | 0.5048 | 0.41 | 449.05 |
| 8 | 0.5035 | 0.4084 | 449.65 |
| 9 | 0.5015 | 0.406 | 450.55 |
| 10 | 0.4993 | 0.4035 | 451.51 |
| 14 | 0.4925 | 0.3921 | 456.57 |

The microbiome disruption half-life with the lowest AIC (i.e. best fit) was used for subsequent analyses.

#### Supplementary Table 2 – Relative abundance of major taxa in baseline samples

| Taxon | Median relative abundance (IQR) % |
|---|---|
| *Enterococcus* | 0.13 (0.056 - 1.3) |
| *Enterobacteriaceae* | 1.4 (0.055 - 7.7) |
| Bacteroidetes | 38 (18 - 61) |
| Clostridia | 22 (10 - 40) |
| Actinobacteria | 3.3 (0.70 - 9.1) |
| *All taxa above* | *91 (85 - 96)* |

### ARMORD Case Report Form

#### Supplementary Information 1 – Case Report Form

### Antibiotic Resistance in the Microbiome OxfoRD (ARMORD) Study – Study Questionnaire for investigator completion

version 3.0 (25th November 2015) Author: N Fawcett

THIS IS A RESEARCH STUDY OFFICIAL DOCUMENT AND IS HIGHLY CONFIDENTIAL. IF YOU ARE NOT A MEMBER OF THE ARMORd RESEARCH TEAM AND HAVE OBTAINED THIS DOCUMENT IN ERROR, PLEASE INFORM THE RESEARCH TEAM IMMEDIATELY ON: /01865 222194, or hand it in to the Microbiology Department on Level 7 of the John Radcliffe Hospital (for attention of Prof Crook Group). We will pick it up and review our handling of this document.

**Please confirm with the participant this statement: "Whilst this information is of great help to us in understanding antibiotic resistance and the microbiome, you are free to choose which questions to answer, and if you require clarification or prefer not to answer, let us know".**

**Study Number:** ____________________ **Date of Questionnaire:** _____________

**Researcher:** _______________________

1) **Tell me about any antibiotics you may have received in the last year:**
Antibiotics in past month? Y / N
Antibiotics in the past year? Y / N
If yes to either then use additional data sheet

2) **Tell me anything you can recall about your antibiotic use throughout your life. For example - we would like to find out if you have a condition like asthma and may have taken many courses of antibiotics every year, or if you are someone who virtually never takes antibiotics.**
Antibiotic use: More than 10 courses in the last 5 years.................[ ]
Antibiotic use: 1 – 10 courses in the last 5 years.................[ ]
Antibiotic use: No antibiotics in the last 5 years.................[ ]
Notes:

3) **Have you had any infections (such as urinary tract infections, pneumonia or skin infections) in the last 3 years?**
Had an infection, think it was resistant.................[ ]
Had an infection, think it was sensitive.................[ ]
Had an infection, but don't know the resistance information.................[ ]
No infection.................[ ]
Details (what do you know about it?):

4) **Tell me about any travel in the last three years:**
No Travel outside UK.................[ ]
Travel outside UK.................[ ] (use additional data sheet)

### ARMORD Case Report Form

**5) What best describes your diet?**

No dietary restrictions......................[ ]
Vegetarian (eats eggs & milk)............[ ]
Vegan (no eggs or milk products)......[ ]

**6) How often do you touch or handle the following food products?**

| | Never | Occasionally/once a month or less | Once a week to once a month | Multiple times a week |
|---|---|---|---|---|
| Raw chicken meat | | | | |
| Other raw meat: | | | | |
| Raw shellfish: (including prawns) | | | | |

**7) Do you work in the food industry?**

No [ ]  Animal farm [ ]  Plant farm [ ]  Abattoir [ ]

**8) Do you work in the healthcare profession?**

No.............................................................................................................[ ]
Hospital, direct physical contact with patients..................................................[ ]
Hospital, no direct contact with patients..........................................................[ ]
Hospital including patient bodily hygiene and contact with waste matter.......[ ]
GP surgery/outpatient clinic* only, direct contact with patients.....................[ ]
GP surgery/clinic*, no direct contact with patients...........................................[ ]
*a healthcare site which doesn't have patients to stay overnight for care

**9) How often have you been to hospital in the last year?**

No contact.....................................................[ ]
As a patient:
Contact, only outpatients/day case................[ ]
Contact, inpatient..........................................[ ]
Last date of contact:

| How much? | 1 day [ ] | 2-14 days [ ] | >14 days [ ] |
|---|---|---|---|

Details:

Do you visit others in hospital regularly?

Notes (place/duration):

**10) Have you been admitted to hospital overnight or more in the last 5 years?**

No [ ]  Yes [ ] (use additional data sheet)

**11) How often have you been in contact with residential/nursing care in the last year?**

As a resident:
Never [ ]  Previous contact, not current [ ]  Current [ ]
Notes (last contact, type of care, total approx. duration):
As a visitor:

| How often as a visitor? | Never | Occasionally/once a month or less | Once a week to once a month | Multiple times a week | Approx. date of last contact |
|---|---|---|---|---|---|
| Nursing Care | | | | | |
| Residential care | | | | | |

**12) How often do you provide childcare for babies or children <2 years?**

| | Never | Occasionally/once a month or less | Once a week to once a month | Multiple times a week | Approx. date of last contact |
|---|---|---|---|---|---|
| Any care | | | | | |
| Care, including personal hygiene for child (body washing and/or handling of nappies) | | | | | |

**13) Do you have any pets in your household?**

Cat.......................[ ]
Dog.......................[ ]
Other(s)................[ ] (please specify):
None.....................[ ]
I clean up their poo Y / N

**14) Have you had any episodes of diarrhoea in the last 2 months?**

No.........................................[ ]
1+ episode of diarrhoea......[ ]
Loose stools on daily basis...[ ]
Notes (cause if known, timing):

**15) Have you been treated with chemotherapy, radiotherapy or medication which affects the immune system* in the last 5 years?**

No.........................[ ]
Yes.......................[ ]
Don't know.............[ ]
Notes (What was it for? How long did you take it for? How did you get given it? e.g. tablets/injections):

* You may have received this if you had an autoimmune disease (overactive immune system) This includes steroids, methotrexate, hydroxychloroquine, sulfasalazine, leflunomide, and injections of anti-TNFa/IL-2R treatment for conditions like rheumatoid arthritis.

**16) Have you taken any of the following in the last month?**

| Medication *(if in doubt , you can discuss at the visit)* | Never | Yes, regularly | Occasionally | Don't know |
|---|---|---|---|---|
| Proton-pump inhibitors (e.g. Omeprazole, lansoprazole) | | | | |
| Non-steroidal anti-inflammatory drugs (e.g. ibuprofen, aspirin, diclofenac, naproxen) | | | | |
| Vitamin, herbal or dietary supplements (including fibre) | | | | |

Notes (What was it called? What was it for? How long did you take it for? How did you get given it? e.g. tablets/injections):

**17) How often do you eat the following foods?**

(if you are tired or short of time, we can leave it or go through this at another visit)

| Type of food (approx. serving size) | Never | Less than once a week | Once a week | 2-4 times a week | 5-6 times a week | Once or more daily | If more than once daily – how many times? |
|---|---|---|---|---|---|---|---|
| High fibre food – brown bread, brown rice, high fibre cereal (one handful/piece of bread) | | | | | | | |
| Other cereals, breads potatoes, rice, pasta (one handful/piece of bread) | | | | | | | |
| Fresh fruit (one apple size) | | | | | | | |
| Fresh vegetables (one apple size) | | | | | | | |
| Milk, cheese and yoghurt products (one glass/ small yoghurt pot) | | | | | | | |
| Meat, fish and poultry (one handful) | | | | | | | |
| Non-animal meat alternatives (Quorn, soy mince) (one handful) | | | | | | | |
| Eggs | | | | | | | |
| Oil and butter (one tablespoon) | | | | | | | |
| Chocolate, cake and sweets (one chocolate bar/handful) | | | | | | | |
| Ready meals/pre-packaged meals | | | | | | | |

**18) Have you ever smoked as much as one cigarette a day for as long as a year?**

No (non-smoker)........................................[ ]

Yes:

I have smoked this amount previously but have not smoked a cigarette in the last year........[ ]

I have smoked less than one cigarette a day in the last year........................................[ ]

I currently smoke one cigarette or more, on average, daily........................................[ ]

Notes:

**19) How many units a week do you drink of alcohol (on average in the last year)?**

(pint of beer = 2 units, small glass of wine = 1 unit, large glass of wine = 2 units, shot of spirits = 1 unit)

Units/week:

Never drink alcohol [ ]

**20) Do you have anything else of relevance you think we should know?**

**ARMORD additional data sheet for investigator (enter approximate values if unknown)**

| Antimicrobial (non-OUH in past year, U = unknown) | Route | Start date | Stop date or duration (d/w/m) | Indication (blank if unknown) |
|---|---|---|---|---|

| Hospital (admissions outside OUH in past 5 years) | Admission | Discharge or duration (d/w/m) | Reason (blank if unknown) |
|---|---|---|---|

| Country (past 3 years, record only last visit to each country) | Start | End or duration (d/w/m) |
|---|---|---|
