## Supplementary material for "The impact of different antimicrobial exposures on the gut microbiome in the ARMORD observational study": data_description.txt

Description of ARMORD data

### Study data
1) AMR_genes_ARIBA_CARD.csv
Output of ARIBA AMR gene local assembler report for each sequenced sample (run using CARD ressitance database). One row per assembled AMR gene. Further details can be found here https://github.com/sanger-pathogens/ariba/wiki/Task%3A-run
pid - participant ID 
no - sample number
samp_id	- sample ID (includes alphanumeric participant ID and sample number)
ref_name - original name of reference sequence chosen from cluster
CARD_accession_number - AMR gene accession number in CARD database
cluster - name of CARD AMR gene cluster
var_only - 1=variant only, 0=presence/absence
rpkm - reads in cluster per AMR gene kilobase per million reads
rpm - reads in cluster per million reads
reads - number of reads
nonhuman_reads - number of nonhuman reads (all subsampled to 3.5 million reads)
max_cov - number of reference nucleotides assembled by this contig divided by reference gene length (i.e. ref_base_assembled/ref_len)
pc_ident - %identity between reference sequence and contig
ref_len - reference gene length
ref_base_assembled - number of reference nucleotides assembled by this contig
ctg_len - length of contig
ctg_cov - mean mapped read depth of this contig
free_text - other free text about reference sequence, from CARD database

2) Antimicrobial_exposures.csv
Antimicrobial data from electronic records and study CRFs, each row is a course of antimicrobials
pid - participant ID 
drug - antimicrobial name
route - route of administration
first_sample - first stool sample from participant (used for relative date of course start/end)
drug_group - category of antimicrobial (A = aminoglycoside, B = beta_lactam_broad, C = clindamycin, F = antifolate, G = glycopeptide, M = macrolide, N = metronidazole, O = other, P = beta_lactam_narrow, Q = quinolone, T = tetracycline, U = unknown, V = antiviral, Z = antifungal)
dose_interval - dosing interval of drug
firstdose_days_after_first_sample - start date of course in days, relative to first stool sample
lastdose_days_after_first_sample - end date of course in days, relative to first stool sample

3) card.obo 
CARD AMR ontology/database file (for reference - was used to create AMR_genes_ARIBA_CARD.csv output but not needed to reproduce plots)

4) patients.csv
List of ARMORD participants with one row per person
pid - participant ID (numeric, note an alphanumeric system was also used in which CS = cross-sectional and LS = longitudinal, both starting from 1, this was replaced with a purely numeric system in which LS patients started from 500)
age_category - ordinal age (5 year groups)
age_group - 5-year age group at enrolment
sex - sex (1 = male, 0 = female)
category - Medical = medical in/outpatient, Healthy = healthy volunteer, Haem_autograft = haematology patient having autologous SCT, Heam_allograft = haematology patient having allogeneic SCT

5) Samples.csv
List of sequenced samples. Not all collected samples were sequenced, so sample numbering for participants is not necessarily consecutive
pid - participant ID 
sample_order - sample number for that participant, starting at 1 for first sample
samp_id - sample ID (combining alphanumeric participant ID and sample order)
collected_days_after_first_sample - time of collection relative to first sample (days)
bristol_score - sample consistency on bristol scale
filtered_reads - number of reads filtered during QC
human_reads - number of human reads in original sequence data (removed prior to analysis & removed from sequence data in public repository)
nonhuman_reads (number of nonhuman reads - subsampled to 3500000)
total_reads (total reads)

6) Taxa_Metaphlan.csv
Taxonomic profile from Metaphlan
samp_id - sample id
kingdom/phylum/class/order/family/genus/species - taxa from metaphlan output
perc - percentage abundance of taxon
seq_run - sequence run


### Figure data
These are outputs of the linear models used to plot figures
