## Supplementary material for "The impact of different antimicrobial exposures on the gut microbiome in the ARMORD observational study": ARMORD R project: run_all.R

```
# Loads primary study data and input data for models
# Plots figures for ARMORD paper (note the public dataset uses 5 year age categories for pseudonymisation, leading to slight difference from results presented in the manuscript)
# Exports figure pdfs in /plots folder, and model outputs in /exports folder
dir.create("exports", showWarnings = F)
dir.create("plots", showWarnings = F)
source(file = "src/1_load_data.R")
source(file = "src/Fig1A_cross_sectional_drug_diversity.R")
source(file = "src/Fig1B_longitudinal_drug_diversity.R")
source(file = "src/Fig2A_cross_sectional_drug_taxa.R")
source(file = "src/Fig2B_longitudinal_drug_taxa.R")
source(file = "src/Fig3A_cross_sectional_drug_ARG.R")
source(file = "src/Fig3B_longitudinal_drug_ARG.R")
source(file = "src/SuppFig1_model_areas.R")
source(file = "src/SuppFig2_model_courses.R")
source(file = "src/SuppFig4A_cross_sectional_class_diversity.R")
source(file = "src/SuppFig4B_longitudinal_class_diversity.R")
source(file = "src/SuppFig5A_cross_sectional_class_taxa.R")
source(file = "src/SuppFig5B_longitudinal_class_taxa.R")
source(file = "src/SuppFig6A_cross_sectional_class_ARG.R")
source(file = "src/SuppFig6B_longitudinal_class_ARG.R")
source(file = "src/SuppData.R")
```
