## Supplementary material for "The impact of different antimicrobial exposures on the gut microbiome in the ARMORD observational study": ARMORD R project: 1_load_data.R

```
## Load libraries & primary data

## load libraries ------------------
library(lmtest)
library(multiwayvcov)
library(tidyverse)
library(stringr)
library(stringi)
library(readr)
library(forcats)
library(scales)
#library(ontologyIndex) ## required if browsing CARD AMR ontology
theme_set(theme_bw())

## load data ------------------
data_folder = paste0(getwd(), "/armord_data/study_data/")

for (file in list.files(data_folder)) {
  if (str_detect(file, ".csv")) {
    name <- stri_replace(file, replacement = "", regex = ".csv")
    assign(name, read_csv(paste0(data_folder, file), guess_max = 10000), globalenv())
}}

## CARD AMR ontology - uncomment if loading CARD ontology for browsing
# a_card <-
#   get_ontology(paste0(data_folder,"card.obo"),
#                propagate_relationships = c("is_a",
#                                            "part_of",
#                                            "confers_resistance_to_drug",
#                                            "confers_resistance_to"))

data_folder = paste0(getwd(), "/armord_data/model_data/")

for (file in list.files(data_folder)) {
  if (str_detect(file, ".csv")) {
    name <- stri_replace(file, replacement = "", regex = ".csv")
    assign(name, read_csv(paste0(data_folder, file), guess_max = 10000), globalenv())
  }}

## remove temp objects ------------------
rm(file, name, data_folder)

# list of plotted antimicrobials
names_of_AM_drug_exposures_excluding_rarities <- c("aciclovir_po", "amoxicillin_iv", "amoxicillin_po", "ceftazidime_iv", "ceftriaxone_iv", "ciprofloxacin_po", "clarithromycin_po", "clindamycin_po", "co_amoxiclav_iv", "co_amoxiclav_po", "co_trimoxazole_po", "doxycycline_po", "flucloxacillin_po", "fluconazole_po", "gentamicin_iv", "meropenem_iv", "metronidazole_iv", "metronidazole_po", "nitrofurantoin_po", "piptaz_iv", "posaconazole_po", "trimethoprim_po", "vancomycin_iv", "voriconazole_po")

names_of_AM_class_exposures_excluding_rarities <- c("aminoglycoside", "antifolate", "antifungal", "antiviral", "beta_lactam_broad", "beta_lactam_narrow", "clindamycin","glycopeptide", "macrolide", "metronidazole", "quinolone", "tetracycline", "unknown")

names_of_pair_AM_drug_exposures_excluding_rarities <- c("aciclovir_iv", "aciclovir_po", "ciprofloxacin_po", "co_amoxiclav_po", "co_trimoxazole_po", "fluconazole_po", "gentamicin_iv", "meropenem_iv", "penicillin_V_po", "pentamidine_iv", "piptaz_iv", "posaconazole_po", "vancomycin_iv", "voriconazole_po")

names_of_pair_AM_class_exposures_excluding_rarities <- c("aminoglycoside", "antifolate", "antifungal", "antiviral", "beta_lactam_broad", "beta_lactam_narrow", "glycopeptide", "quinolone")

# antimicrobial exposure decay half life (days) for illustrative figures - supp figs 1&2 (selected as best fit - see manuscript for details)
decay_half_life <-  6

# function for formatting powers of ten in plots
scientific <- function(x){ifelse(x==0, "0", ifelse(x == 1, "1", parse(text=gsub("[+]", "", gsub("e", "%*%10^", scientific_format()(x))))))}
```
