## Supplementary material for "The impact of different antimicrobial exposures on the gut microbiome in the ARMORD observational study": ARMORD R project: Fig1A_cross_sectional_drug_diversity.R

```
# PLOT FIGURE 1A
# Data = Cross-sectional samples
# Exposure = Antimicrobial drug-route (+ covariates)
# Outcome = Shannon diversity
# Requires output of scripts 1, 2 & 3

### Data table  ----
data_for_CS_AM_drug_diversity_model <- 
  b_first_samples %>%
  select(pid, no, samp_id) %>% 
  left_join(c_patients, "pid") %>% 
  left_join(c_conditioning, c("samp_id")) %>% 
  left_join(c_cat_max_news_pre_sample, c("pid", "samp_id")) %>% 
  left_join(c_cat_charlson, c("pid", "samp_id")) %>% 
  left_join(c_wcc, "samp_id") %>% 
  left_join(c_crp, "samp_id") %>% 
  left_join(table_of_samples_with_AM_drug_exposures, "samp_id") %>%
  left_join(c_diversity_indices_mp, c("pid", "no", "samp_id"))

names_of_AM_drug_exposures_excluding_rarities <- c("aciclovir_po", "amoxicillin_iv", "amoxicillin_po", "ceftazidime_iv", "ceftriaxone_iv", "ciprofloxacin_po", "clarithromycin_po", "clindamycin_po", "co_amoxiclav_iv", "co_amoxiclav_po", "co_trimoxazole_po", "doxycycline_po", "flucloxacillin_po", "fluconazole_po", "gentamicin_iv", "meropenem_iv", "metronidazole_iv", "metronidazole_po", "nitrofurantoin_po", "piptaz_iv", "posaconazole_po", "trimethoprim_po", "vancomycin_iv", "voriconazole_po")

### Exposures ----
names_of_all_exposures_in_CS_AM_drug_diversity_model <- c(
  names_of_AM_drug_exposures_excluding_rarities,
  "age_category",
  "sex",
  "max_charlson",
  "category",
  "max_tt",
  "cat_high_max_wcc",
  "cat_low_min_wcc",
  "cat_high_max_crp",
  "trunc_conditioning_day")

### Diversity model ------------------
multivariable_CS_AM_drug_diversity_model <- 
  lm(as.formula(paste0("shannon ~ ",
                       paste(names_of_all_exposures_in_CS_AM_drug_diversity_model, 
                             collapse = " + "))),
     data = data_for_CS_AM_drug_diversity_model)

multivariable_CS_AM_drug_diversity_model_data_frame <- 
  data_frame(variable = summary(multivariable_CS_AM_drug_diversity_model)$coefficients[-1,2] %>% names(), 
             effect = summary(multivariable_CS_AM_drug_diversity_model)$coefficients[-1,1], 
             se = summary(multivariable_CS_AM_drug_diversity_model)$coefficients[-1,2], 
             ci = 1.96*summary(multivariable_CS_AM_drug_diversity_model)$coefficients[-1,2], 
             t = summary(multivariable_CS_AM_drug_diversity_model)$coefficients[-1,3], 
             p = summary(multivariable_CS_AM_drug_diversity_model)$coefficients[-1,4])

# Univariable estimates (via loop as lm not vectorised)
univariable_CS_AM_drug_diversity_model_data_frame <- data_frame(variable = NA_character_, 
                                         univ_effect = NA_real_, 
                                         univ_se = NA_real_, 
                                         univ_ci = NA_real_)
for(loop_variable in names_of_all_exposures_in_CS_AM_drug_diversity_model) {
  loop_df <- data_frame(variable = loop_variable,
                        univ_effect = summary(lm(as.formula(paste0("shannon ~ ", variable)),
                                                 data = data_for_CS_AM_drug_diversity_model))$coefficients[2,1],
                        univ_se = summary(lm(as.formula(paste0("shannon ~ ", variable)),
                                             data = data_for_CS_AM_drug_diversity_model))$coefficients[2,2],
                        univ_ci = 1.96*univ_se)
  univariable_CS_AM_drug_diversity_model_data_frame <- bind_rows(univariable_CS_AM_drug_diversity_model_data_frame, loop_df)
  assign("univariable_CS_AM_drug_diversity_model_data_frame", univariable_CS_AM_drug_diversity_model_data_frame, envir = globalenv())
}
rm(loop_df, loop_variable)

# Patient category treated differently as three categories won't work in the loop above (for analysis - non antimicrobial categories are not plotted below)
patient_category_CS_AM_drug_diversity_model <-
  summary(lm(shannon ~ category, data = data_for_CS_AM_drug_diversity_model))

univariable_CS_AM_drug_diversity_model_data_frame <-
  univariable_CS_AM_drug_diversity_model_data_frame %>% filter(!is.na(variable)) %>%
  bind_rows(data_frame(variable = c("categoryMedical",
                                    "categoryHaem_autograft",
                                    "categoryHaem_allograft"),
                       univ_effect = patient_category_CS_AM_drug_diversity_model$coefficients[2-3,1],
                       univ_se = patient_category_CS_AM_drug_diversity_model$coefficients[2-3,2],
                       univ_ci = 1.96*univ_se))

# combine multivariable & univariable estimates in same data frame
combined_CS_AM_drug_diversity_model_data_frame <- 
  multivariable_CS_AM_drug_diversity_model_data_frame %>% 
  left_join(univariable_CS_AM_drug_diversity_model_data_frame, "variable") %>% 
  left_join(number_of_first_samples_with_each_AM_drug_exposure, c("variable" = "drug_route")) %>% 
  mutate(variable = str_replace_all(variable, "_", " "))

## > plot diversity ------------------
combined_CS_AM_drug_diversity_model_data_frame %>%
  # remove estimates that are very uncertain (and don't differ from zero)
  filter(!is.na(n), ci < 3) %>%
  mutate(variable = str_to_sentence(variable),
         variable = fct_reorder(variable, desc(effect))) %>% 
  ggplot() +
  geom_point(aes(y = variable, x = effect), position = position_nudge(y = -0.15)) +
  geom_errorbarh(aes(y = variable, xmin = effect - ci, xmax = effect + ci), colour = "grey25", height = 0, position = position_nudge(y = -0.15)) +
  # PLOT UNIVARIABLE ESTIMATES
  geom_point(aes(y = variable, x = univ_effect), colour = "grey", alpha = 0.65, position = position_nudge(y = 0.15)) +
  geom_errorbarh(aes(y = variable, xmin = univ_effect - univ_ci, xmax = univ_effect + univ_ci), colour = "grey", alpha = 0.65, height = 0, position = position_nudge(y = 0.15)) +
  geom_vline(xintercept = 0) +
  #FORMATTING
  geom_text(aes(y = variable, x = -4, label = n)) +
  #geom_text(aes(y = variable, x = 2.5, label = paste0(format(round(effect, 1), nsmall = 1), " (", format(round(effect - ci, digits = 1), nsmall = 1), ", ", format(round(effect + ci, digits = 1), nsmall = 1), ")"))) + #IF INCLUDING EFFECT ESTIMATES & 95% CI ON PLOT
  scale_x_continuous(breaks = c(-3, -2, -1, 0, 1, 2, 3)) +
  coord_cartesian(xlim = c(-4,4)) +
  labs(title = "Figure 1A - Cross-sectional", x = "Change in Shannon diversity", y = "") +
  theme(axis.text.y = element_text(size = 10, face = "bold", colour = "black"),
        axis.text.x = element_text(size = 10, face = "bold", colour = "black"),
        axis.line.x = element_blank(),
        axis.line = element_line(colour = "black"))

ggsave("plots/Figure 1A - Antimicrobial drug vs Shannon diversity in cross-sectional arm.pdf", width = 148, height = 210, units = "mm")

write.csv(combined_CS_AM_drug_diversity_model_data_frame |> 
            mutate(n = if_else(!is.na(n), n, 225),
                   lower_ci = effect - ci,
                   upper_ci = effect + ci) |> 
            select("Variable" = variable, 
                   "Multivariable effect" = effect, 
                   "Multivariable std error" = se,
                   "Multivariable lower 95% CI" = lower_ci,
                   "Multivariable upper 95% CI" = upper_ci,
                   "Multivariable p value" = p, 
                   "Univariable effect" = univ_effect, 
                   "Univariable std error" = univ_se, 
                   "Number exposed" = n), 
          "exports/Figure 1A data - Antimicrobial drug vs Shannon diversity in cross-sectional arm.csv", row.names = F)

multivariable_CS_AM_drug_diversity_model %>% summary()
multivariable_CS_AM_drug_diversity_model %>% AIC()

# remove temporary variables (note combined data frame not removed as needed for longitudinal plot)
rm(#data_for_CS_AM_drug_diversity_model, 
names_of_all_exposures_in_CS_AM_drug_diversity_model, 
multivariable_CS_AM_drug_diversity_model, 
multivariable_CS_AM_drug_diversity_model_data_frame, 
univariable_CS_AM_drug_diversity_model_data_frame, 
patient_category_CS_AM_drug_diversity_model)
```
