## Supplementary material for "The impact of different antimicrobial exposures on the gut microbiome in the ARMORD observational study": ARMORD R project: Fig2B_longitudinal_drug_taxa.R

```
# PLOT FIGURE 2B
# Data = Longitudinal samples
# Exposure = Antimicrobial drug (+ covariates)
# Outcome = Abundance of selected bacterial taxa
# Requires output of scripts 1, 2 & 3

### Data table  ----
data_for_LS_AM_drug_taxa_model <- 
  l_pairs %>% 
  left_join(l_patients %>% select(pid, age_category, sex, tx), "pid") %>% 
  left_join(l_wcc, "pair_id") %>% 
  left_join(l_crp, "pair_id") %>% 
  left_join(l_news, "pair_id") %>% 
  left_join(table_of_pairs_with_AM_drug_exposures, "pair_id") %>%
  left_join(l_bugRA, "pair_id") %>%
  mutate(conditioning_day = collected.y)

### Exposures ----
names_of_all_exposures_in_LS_AM_drug_taxa_model <- c(
  names_of_pair_AM_drug_exposures_excluding_rarities,
  "age_category",
  "sex",
  "tx",
  "conditioning_day",
  "sample_separation",
  "new_low_wcc",
  "new_high_wcc",
  "new_high_crp",
  "news_increase")

### Bug models ----
# Note inclusion of baseline abundance as covariate

# > Enterobateriaceae ----
multivariable_LS_AM_drug_entb_model <- 
  lm(as.formula(paste0("log_entbRA_trunc_diff ~ ", 
                       paste0(names_of_all_exposures_in_LS_AM_drug_taxa_model, collapse = " + "), 
                       " + log_entbRA_trunc.x")),
     data = data_for_LS_AM_drug_taxa_model)

robust_multivariable_LS_AM_drug_entb_model <- 
  coeftest(multivariable_LS_AM_drug_entb_model, 
           cluster.vcov(multivariable_LS_AM_drug_entb_model, data_for_LS_AM_drug_taxa_model$pid))

robust_multivariable_LS_AM_drug_entb_model_data_frame <- 
  data_frame(variable = robust_multivariable_LS_AM_drug_entb_model[-1,2] %>% names(), 
             effect = robust_multivariable_LS_AM_drug_entb_model[-1,1], 
             se = robust_multivariable_LS_AM_drug_entb_model[-1,2], 
             ci = 1.96*robust_multivariable_LS_AM_drug_entb_model[-1,2], 
             t = robust_multivariable_LS_AM_drug_entb_model[-1,3], 
             p = robust_multivariable_LS_AM_drug_entb_model[-1,4]) |> 
  mutate(effect_fold = 10^effect,
         upper = 10^(effect + ci),
         lower = 10^(effect - ci)) %>% 
  mutate(group = "entb")

# > Enterococci ----
multivariable_LS_AM_drug_entc_model <- 
  lm(as.formula(paste0("log_entcRA_trunc_diff ~ ", 
                       paste0(names_of_all_exposures_in_LS_AM_drug_taxa_model, collapse = " + "), 
                       " + log_entcRA_trunc.x")),
     data = data_for_LS_AM_drug_taxa_model)

robust_multivariable_LS_AM_drug_entc_model <- 
  coeftest(multivariable_LS_AM_drug_entc_model, 
           cluster.vcov(multivariable_LS_AM_drug_entc_model, data_for_LS_AM_drug_taxa_model$pid))

robust_multivariable_LS_AM_drug_entc_model_data_frame <- 
  data_frame(variable = robust_multivariable_LS_AM_drug_entc_model[-1,2] %>% names(), 
             effect = robust_multivariable_LS_AM_drug_entc_model[-1,1], 
             se = robust_multivariable_LS_AM_drug_entc_model[-1,2], 
             ci = 1.96*robust_multivariable_LS_AM_drug_entc_model[-1,2], 
             t = robust_multivariable_LS_AM_drug_entc_model[-1,3], 
             p = robust_multivariable_LS_AM_drug_entc_model[-1,4]) |> 
  mutate(effect_fold = 10^effect,
         upper = 10^(effect + ci),
         lower = 10^(effect - ci)) %>% 
  mutate(group = "entc")

# > Bacteroidetes ----
multivariable_LS_AM_drug_bact_model <- 
  lm(as.formula(paste0("log_bactRA_trunc_diff ~ ", 
                       paste0(names_of_all_exposures_in_LS_AM_drug_taxa_model, collapse = " + "), 
                       " + log_bactRA_trunc.x")),
     data = data_for_LS_AM_drug_taxa_model)

robust_multivariable_LS_AM_drug_bact_model <- 
  coeftest(multivariable_LS_AM_drug_bact_model, 
           cluster.vcov(multivariable_LS_AM_drug_bact_model, data_for_LS_AM_drug_taxa_model$pid))

robust_multivariable_LS_AM_drug_bact_model_data_frame <- 
  data_frame(variable = robust_multivariable_LS_AM_drug_bact_model[-1,2] %>% names(), 
             effect = robust_multivariable_LS_AM_drug_bact_model[-1,1], 
             se = robust_multivariable_LS_AM_drug_bact_model[-1,2], 
             ci = 1.96*robust_multivariable_LS_AM_drug_bact_model[-1,2], 
             t = robust_multivariable_LS_AM_drug_bact_model[-1,3], 
             p = robust_multivariable_LS_AM_drug_bact_model[-1,4]) |> 
  mutate(effect_fold = 10^effect,
         upper = 10^(effect + ci),
         lower = 10^(effect - ci)) %>% 
  mutate(group = "bact")

# > Clostridia ----
multivariable_LS_AM_drug_clos_model <- 
  lm(as.formula(paste0("log_closRA_trunc_diff ~ ", 
                       paste0(names_of_all_exposures_in_LS_AM_drug_taxa_model, collapse = " + "), 
                       " + log_closRA_trunc.x")),
     data = data_for_LS_AM_drug_taxa_model)

robust_multivariable_LS_AM_drug_clos_model <- 
  coeftest(multivariable_LS_AM_drug_clos_model, 
           cluster.vcov(multivariable_LS_AM_drug_clos_model, data_for_LS_AM_drug_taxa_model$pid))

robust_multivariable_LS_AM_drug_clos_model_data_frame <- 
  data_frame(variable = robust_multivariable_LS_AM_drug_clos_model[-1,2] %>% names(), 
             effect = robust_multivariable_LS_AM_drug_clos_model[-1,1], 
             se = robust_multivariable_LS_AM_drug_clos_model[-1,2], 
             ci = 1.96*robust_multivariable_LS_AM_drug_clos_model[-1,2], 
             t = robust_multivariable_LS_AM_drug_clos_model[-1,3], 
             p = robust_multivariable_LS_AM_drug_clos_model[-1,4]) |> 
  mutate(effect_fold = 10^effect,
         upper = 10^(effect + ci),
         lower = 10^(effect - ci)) %>% 
  mutate(group = "clos")

# > Actinobacteria  ----
multivariable_LS_AM_drug_acti_model <- 
  lm(as.formula(paste0("log_actiRA_trunc_diff ~ ", 
                       paste0(names_of_all_exposures_in_LS_AM_drug_taxa_model, collapse = " + "), 
                       " + log_actiRA_trunc.x")),
     data = data_for_LS_AM_drug_taxa_model)

robust_multivariable_LS_AM_drug_acti_model <- 
  coeftest(multivariable_LS_AM_drug_acti_model, 
           cluster.vcov(multivariable_LS_AM_drug_acti_model, data_for_LS_AM_drug_taxa_model$pid))

robust_multivariable_LS_AM_drug_acti_model_data_frame <- 
  data_frame(variable = robust_multivariable_LS_AM_drug_acti_model[-1,2] %>% names(), 
             effect = robust_multivariable_LS_AM_drug_acti_model[-1,1], 
             se = robust_multivariable_LS_AM_drug_acti_model[-1,2], 
             ci = 1.96*robust_multivariable_LS_AM_drug_acti_model[-1,2], 
             t = robust_multivariable_LS_AM_drug_acti_model[-1,3], 
             p = robust_multivariable_LS_AM_drug_acti_model[-1,4]) |> 
  mutate(effect_fold = 10^effect,
         upper = 10^(effect + ci),
         lower = 10^(effect - ci)) %>% 
  mutate(group = "acti")

# Merge tables ----
combined_LS_AM_drug_taxa_model_data_frame <-
  bind_rows(robust_multivariable_LS_AM_drug_entb_model_data_frame, 
            robust_multivariable_LS_AM_drug_entc_model_data_frame, 
            robust_multivariable_LS_AM_drug_bact_model_data_frame, 
            robust_multivariable_LS_AM_drug_clos_model_data_frame, 
            robust_multivariable_LS_AM_drug_acti_model_data_frame) %>%
  right_join(number_of_first_samples_with_each_AM_drug_exposure |> select(drug_route) |> 
               full_join(data_frame(group = c("entb", "entc", "bact", "clos", "acti")), by = character()) , 
             c("variable" = "drug_route", "group")) %>% 
  left_join(number_of_pairs_with_each_AM_drug_exposure, c("variable" = "drug_route")) |> 
  filter(variable %in% c(names_of_AM_drug_exposures_excluding_rarities),
         !variable %in% c("unknown", "cefalexin_po", "metronidazole_iv", "metronidazole_po", "trimethoprim_po")) |> 
  mutate(variable = str_replace_all(variable, "_", " "),        
         variable = str_to_sentence(variable),
         variable = fct_reorder(variable, desc(variable)),
         n = if_else(n < 6,"-", as.character(n)))

# Plot ----
ggplot() +
  geom_point(data = combined_LS_AM_drug_taxa_model_data_frame %>% filter(group == "entc"), aes(y = variable, x = effect_fold), position = position_nudge(y = 0.2), colour = "#1b9e77") +
  geom_errorbarh(data = combined_LS_AM_drug_taxa_model_data_frame %>% filter(group == "entc"), aes(y = variable, xmin = lower, xmax = upper), height = 0, position = position_nudge(y = 0.2), colour = "#1b9e77", size = 1) +
  geom_point(data = combined_LS_AM_drug_taxa_model_data_frame %>% filter(group == "entb"), aes(y = variable, x = effect_fold), position = position_nudge(y = 0.1), colour = "#d95f02") +
  geom_errorbarh(data = combined_LS_AM_drug_taxa_model_data_frame %>% filter(group == "entb"), aes(y = variable, xmin = lower, xmax = upper), height = 0, position = position_nudge(y = 0.1), colour = "#d95f02", size = 1) +
  geom_point(data = combined_LS_AM_drug_taxa_model_data_frame %>% filter(group == "bact"), aes(y = variable, x = effect_fold), position = position_nudge(y = 0.0), colour = "#7570b3") +
  geom_errorbarh(data = combined_LS_AM_drug_taxa_model_data_frame %>% filter(group == "bact"), aes(y = variable, xmin = lower, xmax = upper), height = 0, position = position_nudge(y = 0.0), colour = "#7570b3", size = 1) +
  geom_point(data = combined_LS_AM_drug_taxa_model_data_frame %>% filter(group == "clos"), aes(y = variable, x = effect_fold), position = position_nudge(y = -0.1), colour = "#e7298a") +
  geom_errorbarh(data = combined_LS_AM_drug_taxa_model_data_frame %>% filter(group == "clos"), aes(y = variable, xmin = lower, xmax = upper), height = 0, position = position_nudge(y = -0.1), colour = "#e7298a", size = 1) +
  geom_point(data = combined_LS_AM_drug_taxa_model_data_frame %>% filter(group == "acti"), aes(y = variable, x = effect_fold), position = position_nudge(y = -0.2), colour = "#66a61e") +
  geom_errorbarh(data = combined_LS_AM_drug_taxa_model_data_frame %>% filter(group == "acti"), aes(y = variable, xmin = lower, xmax = upper), height = 0, position = position_nudge(y = -0.2), colour = "#66a61e", size = 1) +
  geom_vline(xintercept = 1) +
  geom_text(data = combined_LS_AM_drug_taxa_model_data_frame %>% filter(group == "entc"),
            aes(y = variable,
                x = 10^4.4,
                label = n)) +
  # TAXA LABELS - not needed if on opposing panel
  # geom_label(aes(x = 10^4.5, y = 8.5, label = "Entercoccus faecium"), colour = "#1b9e77", fontface = "bold", hjust = "right") +
  # geom_label(aes(x = 10^4.5, y = 5, label = "Enterobacteriaceae"), colour = "#d95f02", fontface = "bold", hjust = "right") +
  # geom_label(aes(x = 10^4.5, y = 4.5, label = "Bacteroidetes"), colour = "#7570b3", fontface = "bold", hjust = "right") +
  # geom_label(aes(x = 10^4.5, y = 4, label = "Clostridia"), colour = "#e7298a", fontface = "bold", hjust = "right") +
  # geom_label(aes(x = 10^4.5, y = 3.5, label = "Actinobacteria"), colour = "#66a61e", fontface = "bold", hjust = "right") +
  scale_x_log10(breaks = c(1e-4, 1e-3, 1e-2, 1e-1, 1, 1e1, 1e2, 1e3, 1e4), label = scientific) +
  scale_y_discrete(position = "right") +
  coord_cartesian(xlim = c(10^-4.3, 10^4.3)) +
  labs(title = "Figure 2B - Longitudinal", x = "Change in relative abundance", y = "") +
  theme(axis.text.y = element_text(size = 10, face = "bold", colour = "black"),
        axis.text.x = element_text(size = 10, face = "bold", colour = "black"),
        axis.line.x = element_blank(),
        axis.line = element_line(colour = "black"))

ggsave("plots/Figure 2B - Antimicrobial drug vs selected taxa in longitudinal arm.pdf", width = 210, height = 297, units = "mm")

write.csv(combined_LS_AM_drug_taxa_model_data_frame |> 
            select("Variable" = variable, 
                   "Multivariable effect" = effect, 
                   "Multivariable std error" = se, 
                   "Multivariable p value" = p,
                   "Effect multiple" = effect_fold,
                   "Upper 95% CI" = upper,
                   "Lower 95% CI" = lower,
                   "Organism group" = group,
                   "Number exposed" = n), 
          "exports/Figure 2B data - Antimicrobial drug vs selected taxa in longitudinal arm.csv", row.names = F)

# remove temporary variables (note combined data frame not removed as needed for longitudinal plot)
rm(#data_for_LS_AM_drug_taxa_model,
   names_of_all_exposures_in_LS_AM_drug_taxa_model,
   multivariable_LS_AM_drug_entb_model,
   multivariable_LS_AM_drug_entc_model,
   multivariable_LS_AM_drug_bact_model,
   multivariable_LS_AM_drug_clos_model,
   multivariable_LS_AM_drug_acti_model,
   robust_multivariable_LS_AM_drug_entb_model,
   robust_multivariable_LS_AM_drug_entc_model,
   robust_multivariable_LS_AM_drug_bact_model,
   robust_multivariable_LS_AM_drug_clos_model,
   robust_multivariable_LS_AM_drug_acti_model,
   robust_multivariable_LS_AM_drug_entb_model_data_frame,
   robust_multivariable_LS_AM_drug_entc_model_data_frame,
   robust_multivariable_LS_AM_drug_bact_model_data_frame,
   robust_multivariable_LS_AM_drug_clos_model_data_frame,
   robust_multivariable_LS_AM_drug_acti_model_data_frame)
```
