## Supplementary material for "The impact of different antimicrobial exposures on the gut microbiome in the ARMORD observational study": ARMORD R project: Fig3A_cross_sectional_drug_ARG.R

```
# PLOT FIGURE 3A
# Data = Cross-sectional samples
# Exposure = Antimicrobial group (+ covariates)
# Outcome = Abundance of selected bacterial AMR genes
# Requires output of scripts 1, 2 & 3

### Data table  ----
data_for_CS_AM_drug_ARG_model <- 
  b_first_samples %>%
  select(pid, no, samp_id) %>% 
  left_join(c_patients, "pid") %>% 
  left_join(c_conditioning, c("samp_id")) %>% 
  left_join(c_cat_max_news_pre_sample, c("pid", "samp_id")) %>% 
  left_join(c_cat_charlson, c("pid", "samp_id")) %>% 
  left_join(c_wcc, "samp_id") %>% 
  left_join(c_crp, "samp_id") %>% 
  left_join(table_of_samples_with_AM_drug_exposures, "samp_id") %>% 
  left_join(c_argRA, c("pid", "no", "samp_id"))

### Exposures ----
names_of_all_exposures_in_CS_AM_drug_ARG_model <- c(
  names_of_AM_drug_exposures_excluding_rarities,
  "age_category",
  "sex",
  "max_charlson",
  "category",
  "max_tt",
  "cat_high_max_wcc",
  "cat_low_min_wcc",
  "cat_high_max_crp",
  "trunc_conditioning_day")

### ARG models ------------------
# > Beta-lactamase ----
multivariable_CS_AM_drug_bla_model <- 
  lm(as.formula(paste0("log_bla_rpm_trunc ~ ", 
                       paste(names_of_all_exposures_in_CS_AM_drug_ARG_model, collapse = " + "))),
     data = data_for_CS_AM_drug_ARG_model)

multivariable_CS_AM_drug_bla_model_data_frame <- 
  data_frame(variable = summary(multivariable_CS_AM_drug_bla_model)$coefficients[-1,2] %>% names(), 
             effect = summary(multivariable_CS_AM_drug_bla_model)$coefficients[-1,1], 
             se = summary(multivariable_CS_AM_drug_bla_model)$coefficients[-1,2], 
             ci = 1.96*se, 
             t = summary(multivariable_CS_AM_drug_bla_model)$coefficients[-1,3], 
             p = summary(multivariable_CS_AM_drug_bla_model)$coefficients[-1,4]) %>% 
  mutate(effect_fold = 10^effect,
         upper = 10^(effect + ci),
         lower = 10^(effect - ci)) %>% 
  mutate(group = "bla")

# > Tetracycline (RPP) ----
multivariable_CS_AM_drug_tet_model <- 
  lm(as.formula(paste0("log_tet_rpm_trunc ~ ", 
                       paste(names_of_all_exposures_in_CS_AM_drug_ARG_model, collapse = " + "))),
     data = data_for_CS_AM_drug_ARG_model)

multivariable_CS_AM_drug_tet_model_data_frame <- 
  data_frame(variable = summary(multivariable_CS_AM_drug_tet_model)$coefficients[-1,2] %>% names(), 
             effect = summary(multivariable_CS_AM_drug_tet_model)$coefficients[-1,1], 
             se = summary(multivariable_CS_AM_drug_tet_model)$coefficients[-1,2], 
             ci = 1.96*se, 
             t = summary(multivariable_CS_AM_drug_tet_model)$coefficients[-1,3], 
             p = summary(multivariable_CS_AM_drug_tet_model)$coefficients[-1,4]) %>% 
  mutate(effect_fold = 10^effect,
         upper = 10^(effect + ci),
         lower = 10^(effect - ci)) %>% 
  mutate(group = "tet")

# > Aminoglycoside (AAC, ANT, APH) ----
multivariable_CS_AM_drug_amg_model <- 
  lm(as.formula(paste0("log_amg_rpm_trunc ~ ", 
                       paste(names_of_all_exposures_in_CS_AM_drug_ARG_model, collapse = " + "))),
     data = data_for_CS_AM_drug_ARG_model)

multivariable_CS_AM_drug_amg_model_data_frame <- 
  data_frame(variable = summary(multivariable_CS_AM_drug_amg_model)$coefficients[-1,2] %>% names(), 
             effect = summary(multivariable_CS_AM_drug_amg_model)$coefficients[-1,1], 
             se = summary(multivariable_CS_AM_drug_amg_model)$coefficients[-1,2], 
             ci = 1.96*se, 
             t = summary(multivariable_CS_AM_drug_amg_model)$coefficients[-1,3], 
             p = summary(multivariable_CS_AM_drug_amg_model)$coefficients[-1,4]) %>% 
  mutate(effect_fold = 10^effect,
         upper = 10^(effect + ci),
         lower = 10^(effect - ci)) %>% 
  mutate(group = "amg")

# > Macrolide (mef & erm) ----
multivariable_CS_AM_drug_mac_model <- 
  lm(as.formula(paste0("log_mac_rpm_trunc ~ ", 
                       paste(names_of_all_exposures_in_CS_AM_drug_ARG_model, collapse = " + "))),
     data = data_for_CS_AM_drug_ARG_model)

multivariable_CS_AM_drug_mac_model_data_frame <- 
  data_frame(variable = summary(multivariable_CS_AM_drug_mac_model)$coefficients[-1,2] %>% names(), 
             effect = summary(multivariable_CS_AM_drug_mac_model)$coefficients[-1,1], 
             se = summary(multivariable_CS_AM_drug_mac_model)$coefficients[-1,2], 
             ci = 1.96*se, 
             t = summary(multivariable_CS_AM_drug_mac_model)$coefficients[-1,3], 
             p = summary(multivariable_CS_AM_drug_mac_model)$coefficients[-1,4]) %>% 
  mutate(effect_fold = 10^effect,
         upper = 10^(effect + ci),
         lower = 10^(effect - ci)) %>% 
  mutate(group = "mac")

# > Glycopeptide (VanA)  ----
multivariable_CS_AM_drug_van_model <- 
  lm(as.formula(paste0("log_van_rpm_trunc ~ ", 
                       paste(names_of_all_exposures_in_CS_AM_drug_ARG_model, collapse = " + "))),
     data = data_for_CS_AM_drug_ARG_model)

multivariable_CS_AM_drug_van_model_data_frame <- 
  data_frame(variable = summary(multivariable_CS_AM_drug_van_model)$coefficients[-1,2] %>% names(), 
             effect = summary(multivariable_CS_AM_drug_van_model)$coefficients[-1,1], 
             se = summary(multivariable_CS_AM_drug_van_model)$coefficients[-1,2], 
             ci = 1.96*se, 
             t = summary(multivariable_CS_AM_drug_van_model)$coefficients[-1,3], 
             p = summary(multivariable_CS_AM_drug_van_model)$coefficients[-1,4]) %>% 
  mutate(effect_fold = 10^effect,
         upper = 10^(effect + ci),
         lower = 10^(effect - ci)) %>% 
  mutate(group = "van")

# Merge tables ----
combined_CS_AM_drug_ARG_model_data_frame <-
  bind_rows(multivariable_CS_AM_drug_bla_model_data_frame, 
            multivariable_CS_AM_drug_tet_model_data_frame, 
            multivariable_CS_AM_drug_amg_model_data_frame, 
            multivariable_CS_AM_drug_mac_model_data_frame, 
            multivariable_CS_AM_drug_van_model_data_frame) %>%
  left_join(number_of_first_samples_with_each_AM_drug_exposure, c("variable" = "drug_route")) %>% 
  mutate(variable = str_replace_all(variable, "_", " "),
         variable = str_to_sentence(variable),
         variable = fct_reorder(variable, desc(variable))) %>% 
  filter(!is.na(n),
         !variable %in% c("Unknown", "Cefalexin po", "Metronidazole iv", "Metronidazole po", "Trimethoprim po")) 

# Plot ----
ggplot() +
  geom_point(data = combined_CS_AM_drug_ARG_model_data_frame %>% filter(group == "tet"), aes(y = variable, x = effect_fold), position = position_nudge(y = 0.2), colour = "#1b9e77") +
  geom_errorbarh(data = combined_CS_AM_drug_ARG_model_data_frame %>% filter(group == "tet"), aes(y = variable, xmin = lower, xmax = upper), height = 0, position = position_nudge(y = 0.2), colour = "#1b9e77", size = 1) +
  geom_point(data = combined_CS_AM_drug_ARG_model_data_frame %>% filter(group == "bla"), aes(y = variable, x = effect_fold), position = position_nudge(y = 0.1), colour = "#d95f02") +
  geom_errorbarh(data = combined_CS_AM_drug_ARG_model_data_frame %>% filter(group == "bla"), aes(y = variable, xmin = lower, xmax = upper), height = 0, position = position_nudge(y = 0.1), colour = "#d95f02", size = 1) +
  geom_point(data = combined_CS_AM_drug_ARG_model_data_frame %>% filter(group == "amg"), aes(y = variable, x = effect_fold), position = position_nudge(y = 0.0), colour = "#7570b3") +
  geom_errorbarh(data = combined_CS_AM_drug_ARG_model_data_frame %>% filter(group == "amg"), aes(y = variable, xmin = lower, xmax = upper), height = 0, position = position_nudge(y = 0.0), colour = "#7570b3", size = 1) +
  geom_point(data = combined_CS_AM_drug_ARG_model_data_frame %>% filter(group == "mac"), aes(y = variable, x = effect_fold), position = position_nudge(y = -0.1), colour = "#e7298a") +
  geom_errorbarh(data = combined_CS_AM_drug_ARG_model_data_frame %>% filter(group == "mac"), aes(y = variable, xmin = lower, xmax = upper), height = 0, position = position_nudge(y = -0.1), colour = "#e7298a", size = 1) +
  geom_point(data = combined_CS_AM_drug_ARG_model_data_frame %>% filter(group == "van"), aes(y = variable, x = effect_fold), position = position_nudge(y = -0.2), colour = "#66a61e") +
  geom_errorbarh(data = combined_CS_AM_drug_ARG_model_data_frame %>% filter(group == "van"), aes(y = variable, xmin = lower, xmax = upper), height = 0, position = position_nudge(y = -0.2), colour = "#66a61e", size = 1) +
  geom_vline(xintercept = 1) +
  geom_text(data = combined_CS_AM_drug_ARG_model_data_frame %>% filter(group == "tet"),
            aes(y = variable,
                x = 10^-4.4,
                label = n)) +
  geom_label(aes(x = 10^4.65, y = 12.6, label = "Tetracycline (RPP)"), colour = "#1b9e77", fontface = "bold", hjust = "right", size = 3.5) +
  geom_label(aes(x = 10^4.65, y = 11.9, label = "Beta-lactamase\n(CTX-M/OXA/TEM/SHV)"), colour = "#d95f02", fontface = "bold", hjust = "right", size = 3.5) +
  geom_label(aes(x = 10^4.65, y = 11, label = "Aminoglycoside\n(AAC/ANT/APH)"), colour = "#7570b3", fontface = "bold", hjust = "right", size = 3.5) +
  geom_label(aes(x = 10^4.65, y = 10.1, label = "Macrolide/Clindamycin\n(mef/erm)"), colour = "#e7298a", fontface = "bold", hjust = "right", size = 3.5) +
  geom_label(aes(x = 10^4.65, y = 9.4, label = "Glycopeptide (VanA)"), colour = "#66a61e", fontface = "bold", hjust = "right", size = 3.5) +
  scale_x_log10(breaks = c(1e-4, 1e-3, 1e-2, 1e-1, 1, 1e1, 1e2, 1e3, 1e4), label = scientific) +
  coord_cartesian(xlim = c(10^-4.3, 10^4.3)) +
  labs(title = "Figure 3A - Cross-sectional", x = "Change in relative abundance", y = "") +
  theme(axis.text.y = element_text(size = 10, face = "bold", colour = "black"),
        axis.text.x = element_text(size = 10, face = "bold", colour = "black"),
        #panel.border = element_blank(),
        axis.line.x = element_blank(),
        axis.line = element_line(colour = "black"))

ggsave("plots/Figure 3A - Antimicrobial drug vs selected AMR genes in cross-sectional arm.pdf", width = 210, height = 297, units = "mm")

write.csv(combined_CS_AM_drug_ARG_model_data_frame |> 
            select("Variable" = variable, 
                   "Multivariable effect" = effect, 
                   "Multivariable std error" = se, 
                   "Multivariable p value" = p,
                   "Effect multiple" = effect_fold,
                   "Upper 95% CI" = upper,
                   "Lower 95% CI" = lower,
                   "ARG group" = group,
                   "Number exposed" = n), 
          "exports/Figure 3A data - Antimicrobial drug vs selected AMR genes in cross-sectional arm.csv", row.names = F)

# remove temporary variables (note combined data frame not removed as needed for longitudinal plot)
rm(#data_for_CS_AM_drug_ARG_model,
   names_of_all_exposures_in_CS_AM_drug_ARG_model,
   multivariable_CS_AM_drug_bla_model,
   multivariable_CS_AM_drug_tet_model,
   multivariable_CS_AM_drug_amg_model,
   multivariable_CS_AM_drug_mac_model,
   multivariable_CS_AM_drug_van_model,
   multivariable_CS_AM_drug_bla_model_data_frame,
   multivariable_CS_AM_drug_tet_model_data_frame,
   multivariable_CS_AM_drug_amg_model_data_frame,
   multivariable_CS_AM_drug_mac_model_data_frame,
   multivariable_CS_AM_drug_van_model_data_frame)
```
