## Supplementary material for "The impact of different antimicrobial exposures on the gut microbiome in the ARMORD observational study": ARMORD R project: SuppData.R

```
# Create combined tables with model input data for cross-sectional and logitudinal data
# requires 'data for x model' to be retained not deleted in each script

## Cross-sectional model data -----
cross_sectional_model_input_data <- 
  data_for_CS_AM_drug_diversity_model |> 
  left_join(data_for_CS_AM_drug_taxa_model |> 
              select(pid,
                     "log_enterobacteriaceae_relative_abundance" = log_entbRA_trunc, 
                     "log_enterococcus_relative_abundance" = log_entcRA_trunc, 
                     "log_bacteroidetes_relative_abundance" = log_bactRA_trunc, 
                     "log_clostridia_relative_abundance" = log_closRA_trunc,
                     "log_actinobacteria_relative_abundance" = log_actiRA_trunc), 
            "pid") |> 
  left_join(data_for_CS_AM_drug_ARG_model |> 
              select(pid, 
                     "log_beta_lactam_ARG_reads_per_million" = log_bla_rpm_trunc, 
                     "log_tetracycline_ARG_reads_per_million" = log_tet_rpm_trunc, 
                     "log_aminoglycoside_ARG_reads_per_million" = log_amg_rpm_trunc, 
                     "log_macrolide_ARG_reads_per_million" = log_mac_rpm_trunc, 
                     "log_glycopeptide_ARG_reads_per_million" = log_van_rpm_trunc), "pid") |> 
  left_join(data_for_CS_AM_class_diversity_model |> 
              select(pid,
                     "aminoglycoside_class" = aminoglycoside,
                     "antifolate_class" =	antifolate,
                     "antifungal_class" =	antifungal,
                     "antiviral_class"	= antiviral,
                     "beta_lactam_broad_class"	= beta_lactam_broad,
                     "beta_lactam_narrow_class"	= beta_lactam_narrow,
                     "clindamycin_class"	= clindamycin,
                     "glycopeptide_class"	= glycopeptide,
                     "macrolide_class"	= macrolide,
                     "metronidazole_class"	= metronidazole,
                     "other_class"	= other,
                     "quinolone_class"	= quinolone,
                     "tetracycline_class"	= tetracycline,
                     "unknown_class"	= unknown), "pid") |>
  rename("patient_id" = pid,
         "truncated_conditioning_day" = trunc_conditioning_day,
         "maximum_charlson_score" = max_charlson,
         "high_max_wcc" = cat_high_max_wcc,
         "low_min_wcc" = cat_low_min_wcc,
         "high_max_crp" = cat_high_max_crp,
         "species_richness" = richness)
  
write.csv(cross_sectional_model_input_data,
          "exports/Cross-sectional model input data.csv", row.names = F)

## Longitudinal model data -----
longitudinal_model_input_data <- 
  data_for_LS_AM_drug_diversity_model |> 
  left_join(data_for_LS_AM_drug_taxa_model |> 
              select(pair_id, 
                     "log_enterobacteriaceae_relative_abundance_baseline" = log_entbRA_trunc.x, 
                     "log_enterococcus_relative_abundance_baseline" = log_entcRA_trunc.x, 
                     "log_bacteroidetes_relative_abundance_baseline" = log_bactRA_trunc.x, 
                     "log_clostridia_relative_abundance_baseline" = log_closRA_trunc.x,
                     "log_actinobacteria_relative_abundance_baseline" = log_actiRA_trunc.x,
                     "log_enterobacteriaceae_relative_abundance_change" = log_entbRA_trunc_diff, 
                     "log_enterococcus_relative_abundance_change" = log_entcRA_trunc_diff, 
                     "log_bacteroidetes_relative_abundance_change" = log_bactRA_trunc_diff, 
                     "log_clostridia_relative_abundance_change" = log_closRA_trunc_diff,
                     "log_actinobacteria_relative_abundance_change" = log_actiRA_trunc_diff
                     ), "pair_id") |> 
  left_join(data_for_LS_AM_drug_ARG_model |> 
              select(pair_id, 
                     "log_beta_lactam_ARG_reads_per_million_baseline" = log_bla_rpm_trunc.x, 
                     "log_tetracycline_ARG_reads_per_million_baseline" = log_tet_rpm_trunc.x, 
                     "log_aminoglycoside_ARG_reads_per_million_baseline" = log_amg_rpm_trunc.x, 
                     "log_macrolide_ARG_reads_per_million_baseline" = log_mac_rpm_trunc.x, 
                     "log_glycopeptide_ARG_reads_per_million_baseline" = log_van_rpm_trunc.x,
                     "log_beta_lactam_ARG_reads_per_million_change" = log_bla_rpm_trunc_diff, 
                     "log_tetracycline_ARG_reads_per_million_change" = log_tet_rpm_trunc_diff, 
                     "log_aminoglycoside_ARG_reads_per_million_change" = log_amg_rpm_trunc_diff, 
                     "log_macrolide_ARG_reads_per_million_change" = log_mac_rpm_trunc_diff, 
                     "log_glycopeptide_ARG_reads_per_million_change" = log_van_rpm_trunc_diff
                     ), "pair_id") |> 
  left_join(data_for_LS_AM_class_diversity_model |> 
              select(pair_id,
                     "aminoglycoside_class" = aminoglycoside,
                     "antifolate_class" =	antifolate,
                     "antifungal_class" =	antifungal,
                     "antiviral_class"	= antiviral,
                     "beta_lactam_broad_class"	= beta_lactam_broad,
                     "beta_lactam_narrow_class"	= beta_lactam_narrow,
                     "clindamycin_class"	= clindamycin,
                     "glycopeptide_class"	= glycopeptide,
                     "macrolide_class"	= macrolide,
                     "metronidazole_class"	= metronidazole,
                     "other_class"	= other,
                     "quinolone_class"	= quinolone,
                     "tetracycline_class"	= tetracycline,
                     "unknown_class"	= unknown), "pair_id")

write.csv(longitudinal_model_input_data,
          "exports/Longitudinal model input data.csv", row.names = F)
```
