## Supplementary material for "The impact of different antimicrobial exposures on the gut microbiome in the ARMORD observational study": ARMORD R project: SuppFig1_model_areas.R

```
data <- data_frame(x = -0.1*0:240, y = 2^-(-x/decay_half_life))

ggplot() +
  geom_line(data = data, aes(x,y)) +
  geom_ribbon(data = subset(data, x>=-4 & x<=0), aes(x=x, ymax=y), ymin = 0, fill = "red", alpha = 0.5) +
  geom_ribbon(data = subset(data, x>=-15 & x<=-10), aes(x=x, ymax=y), ymin = 0, fill = "red", alpha = 0.5) +  
  geom_ribbon(data = subset(data, x>=-12 & x<=-6), aes(x=x, ymax=y), ymin = 0, fill = "darkgreen", alpha = 0.5) +
  geom_ribbon(data = subset(data, x>=-21 & x<=-16), aes(x=x, ymax=y), ymin = 0, fill = "blue", alpha = 0.5) +
  geom_segment(aes(x = 0, xend = 0, y = 0, yend = 1), colour = "black") +
  geom_segment(aes(x = 0, xend = -24, y = 0), yend = 0) +
  geom_segment(aes(x = -4, xend = 0, y = 1.05, yend = 1.05), colour = "red", size = 3, alpha = 0.5) +
  geom_text(aes(x = -2, y = 1.08, label = "Antimicrobial B")) +
  geom_segment(aes(x = -15, xend = -10, y = 1.05, yend = 1.05), colour = "red", size = 3, alpha = 0.5) +
  geom_text(aes(x = -12.5, y = 1.08, label = "Antimicrobial B")) +
  geom_segment(aes(x = -12, xend = -6, y = 1.12, yend = 1.12), colour = "darkgreen", size = 3, alpha = 0.5) +
  geom_text(aes(x = -9, y = 1.15, label = "Antimicrobial C")) +
  geom_segment(aes(x = -21, xend = -16, y = 1.12, yend = 1.12), colour = "blue", size = 3, alpha = 0.5) +
  geom_text(aes(x = -18.5, y = 1.15, label = "Antimicrobial A")) +
  geom_segment(aes(x = 0, xend = 0, y = 1.2, yend = 1.1), arrow = arrow(length = unit(0.25, "cm"))) +
  geom_text(aes(x = 0, y = 1.25, label = "Sample\ncollected"), size = 3) + 
  scale_x_continuous(breaks = -28:0, minor_breaks = NULL, labels = 28:0, name = "Days before sample collected", limits = c(-24,0)) +
  scale_y_continuous(breaks = 0:8/8, minor_breaks = NULL, name = NULL) +
  theme(axis.text.y = element_blank(),
        axis.line.y = element_blank(),
        axis.ticks.y = element_blank(),
        panel.border = element_blank())

ggsave("plots/Supplementary Figure 1 - Modelled antimicrobial exposure by course recency.pdf", width = 210, height = 148, units = "mm")
```
