## Supplementary material for "The impact of different antimicrobial exposures on the gut microbiome in the ARMORD observational study": ARMORD R project: SuppFig2_model_courses.R

```
integrand <- function(x) {2^-x}

data_frame(collection_date = 0.1*0:700) %>% # sample collection date (arbitrary day number, starting before exposure)
  full_join(data_frame(course_duration = c(3,7, 14, 28, 42, 100), course_duration_name = c("3 days","7 days", "14 days", "28 days", "42 days", "Continuous")), by = character()) %>% 
  mutate(start_day = 0,
         stop_day = start_day + course_duration,
         relative_start = if_else(collection_date >start_day, collection_date-start_day, 0), # how long ago exposure started (on day 1)
         relative_end = if_else(collection_date >stop_day, collection_date-stop_day,0), # how long ago exposure ended (on day 8)
         area_to_start = sapply(relative_start/decay_half_life, function(x) integrate(integrand, lower = 0, upper = x)$value),
         area_to_end = sapply(relative_end/decay_half_life, function(x) integrate(integrand, lower = 0, upper = x)$value),
         exposure = log(2)*(area_to_start - area_to_end), # multiply area under curve by log(2) so exposures are in the range 0-1
         exposure = if_else(exposure <0.001, 0, exposure),# convert exposures <0.01 to 0, to avoid counting trivial exposures
         course_duration_name = fct_reorder(course_duration_name, course_duration)) %>% 
  ggplot() +
  geom_line(aes(collection_date, exposure, colour = course_duration_name)) +
  geom_segment(aes(x = -2, xend = start_day, y = 0, yend = 0), colour = "#e6ab02") +
  geom_segment(aes(x = start_day, xend = start_day, y = 0.3, yend = 0.2), arrow = arrow(length = unit(0.25, "cm"))) + 
  geom_text(aes(x = start_day, y = 0.4, label = "Course\nstart"), size = 3) + 
  scale_x_continuous(breaks = 7*-1:10, minor_breaks = -7:70, name = "Day of sampling") +
  scale_y_continuous(breaks = 0.1*0:10, name = "Modelled exposure") +
  scale_color_brewer(type = "qual", palette = 2, name = "Course\nduration")

ggsave("plots/Supplementary Figure 2 - Modelled antimicrobial exposure by course duration.pdf", width = 300, height = 100, units = "mm")
```
