## Supplementary material for "The impact of different antimicrobial exposures on the gut microbiome in the ARMORD observational study": ARMORD R project: SuppFig4B_longitudinal_class_diversity.R

```
# PLOT SUPPLEMENTARY FIGURE 4B
# Data = Longitudinal samples
# Exposure = Antimicrobial class (+ covariates)
# Outcome = Shannon diversity
# Requires output of scripts 1, 2, 3 & Figure 1a

### Data table  ----
data_for_LS_AM_class_diversity_model <- 
  l_pairs %>% 
  left_join(l_patients %>% select(pid, age_category, sex, tx), "pid") %>% 
  left_join(l_wcc, "pair_id") %>% 
  left_join(l_crp, "pair_id") %>% 
  left_join(l_news, "pair_id") %>% 
  left_join(table_of_pairs_with_AM_class_exposures, "pair_id") %>%
  left_join(l_diversity_indices, "pair_id") %>%
  mutate(baseline_diversity = shannon.x,
         conditioning_day = collected.y)

### Exposures ----
names_of_all_exposures_in_LS_AM_class_diversity_model <- c(
  names_of_pair_AM_class_exposures_excluding_rarities,
  "age_category",
  "sex",
  "tx",
  "baseline_diversity",
  "conditioning_day",
  "sample_separation",
  "new_low_wcc",
  "new_high_wcc",
  "new_high_crp",
  "news_increase")

### Diversity model ----
multivariable_LS_AM_class_diversity_model <- 
  lm(as.formula(paste0("shannon_diff ~ ", 
                       paste0(names_of_all_exposures_in_LS_AM_class_diversity_model, collapse = " + "))),
     data = data_for_LS_AM_class_diversity_model)

### Robust diversity model ----
# linear model with robust standard errors
robust_multivariable_LS_AM_class_diversity_model <- 
  coeftest(multivariable_LS_AM_class_diversity_model, 
           cluster.vcov(multivariable_LS_AM_class_diversity_model, data_for_LS_AM_class_diversity_model$pid))

robust_multivariable_LS_AM_class_diversity_model_data_frame <- 
  data_frame(variable = robust_multivariable_LS_AM_class_diversity_model[-1,2] %>% names(), 
             effect = robust_multivariable_LS_AM_class_diversity_model[-1,1], 
             se = robust_multivariable_LS_AM_class_diversity_model[-1,2], 
             ci = 1.96*robust_multivariable_LS_AM_class_diversity_model[-1,2], 
             t = robust_multivariable_LS_AM_class_diversity_model[-1,3], 
             p = robust_multivariable_LS_AM_class_diversity_model[-1,4])

# Univariable estimates (via loop as lm not vectorised)
univariable_LS_AM_class_diversity_model_data_frame <- 
  data_frame(variable = NA_character_, 
             univ_effect = NA_real_, 
             univ_se = NA_real_, 
             univ_ci = NA_real_)

for(loop_variable in names_of_all_exposures_in_LS_AM_class_diversity_model) {
  loop_df <- data_frame(variable = loop_variable,
                        univ_effect = coeftest(lm(as.formula(paste0("shannon_diff ~ ", variable)), 
                                                  data = data_for_LS_AM_class_diversity_model), 
                                               cluster.vcov(multivariable_LS_AM_class_diversity_model, 
                                                            data_for_LS_AM_class_diversity_model$pid))[2,1],
                        univ_se = coeftest(lm(as.formula(paste0("shannon_diff ~ ", variable)), 
                                              data = data_for_LS_AM_class_diversity_model), 
                                           cluster.vcov(multivariable_LS_AM_class_diversity_model, 
                                                        data_for_LS_AM_class_diversity_model$pid))[2,2],
                        univ_ci = 1.96*univ_se)
  univariable_LS_AM_class_diversity_model_data_frame <- bind_rows(univariable_LS_AM_class_diversity_model_data_frame, loop_df)
  assign("univariable_LS_AM_class_diversity_model_data_frame", univariable_LS_AM_class_diversity_model_data_frame, envir = globalenv())
}

rm(loop_df, loop_variable)

patient_category_LS_AM_class_diversity_model <- 
  coeftest(lm(shannon_diff ~ tx, 
              data = data_for_LS_AM_class_diversity_model), 
           cluster.vcov(lm(shannon_diff ~ tx, data = data_for_LS_AM_class_diversity_model), 
                        data_for_LS_AM_class_diversity_model$pid))

univariable_LS_AM_class_diversity_model_data_frame <- 
  univariable_LS_AM_class_diversity_model_data_frame %>% filter(!is.na(variable)) %>% 
  bind_rows(data_frame(variable = "txauto", 
                       univ_effect = patient_category_LS_AM_class_diversity_model[2,1],
                       univ_se = patient_category_LS_AM_class_diversity_model[2,2],
                       univ_ci = 1.96*univ_se))

# combine multivariable & univariable estimates in same df
combined_LS_AM_class_diversity_model_data_frame <- 
  robust_multivariable_LS_AM_class_diversity_model_data_frame %>% 
  left_join(univariable_LS_AM_class_diversity_model_data_frame, "variable") %>% 
  left_join(number_of_pairs_with_each_AM_class_exposure, c("variable" = "drug_group_long")) %>% 
  mutate(variable = str_replace_all(variable, "_", " "))

## > plot robust diversity ----
# Note it requires data frame from CS study to plot in consistent format
combined_CS_AM_class_diversity_model_data_frame %>%
  # remove estimates that are very uncertain
  filter(!is.na(n), variable != "unknown") %>% 
  select(variable, cross_sectional_effect = effect) %>% 
  left_join(combined_LS_AM_class_diversity_model_data_frame, "variable") %>%
  mutate(variable = str_to_sentence(variable, locale = "en"),
         variable = fct_reorder(variable, desc(cross_sectional_effect)),
         n = if_else(is.na(n), "-", as.character(n))) %>% 
  ggplot() +
  # PLOT MULTIVARIABLE ESTIMATES
  geom_point(aes(y = variable, x = effect), position = position_nudge(y = -0.15)) +
  geom_errorbarh(aes(y = variable, xmin = effect - ci, xmax = effect + ci), 
                 colour = "grey25", height = 0, position = position_nudge(y = -0.15)) +
  # PLOT UNIVARIABLE ESTIMATES
  geom_point(aes(y = variable, x = univ_effect), colour = "grey", alpha = 0.65, position = position_nudge(y = 0.15)) +
  geom_errorbarh(aes(y = variable, xmin = univ_effect - univ_ci, xmax = univ_effect + univ_ci), 
                 colour = "grey", alpha = 0.65, height = 0, position = position_nudge(y = 0.15)) +
  geom_vline(xintercept = 0) +
  geom_text(aes(y = variable, 
                x = 4, 
                label = n)) +
  ## INCLUDE EFFCT ESTIMATES & 95% CI on plot
  # geom_text(aes(y = variable, x = 3.5, label = paste0(format(round(effect, 1), nsmall = 1), " (", format(round(effect - ci, digits = 1), nsmall = 1), ", ", format(round(effect + ci, digits = 1), nsmall = 1), ")"))) +
  scale_x_continuous(breaks = c(-3, -2, -1, 0, 1, 2, 3)) +
  scale_y_discrete(position = "right") +
  coord_cartesian(xlim = c(-4,4)) +
  labs(title = "Supplementary Figure 4B - Longitudinal", x = "Change in Shannon diversity", y = "") +
  theme(axis.text.y = element_text(size = 10, face = "bold", colour = "black"),
        axis.text.x = element_text(size = 10, face = "bold", colour = "black"),
        axis.line.x = element_blank(),
        axis.line = element_line(colour = "black"))

ggsave("plots/Supplementary Figure 4B - Antimicrobial class vs Shannon diversity in longitudinal arm.pdf", width = 148, height = 210, units = "mm")

write.csv(combined_LS_AM_class_diversity_model_data_frame|> 
            mutate(n = if_else(!is.na(n), n, 173),
                   lower_ci = effect - ci,
                   upper_ci = effect + ci) |> 
            select("Variable" = variable, 
                   "Multivariable effect" = effect, 
                   "Multivariable std error" = se,
                   "Multivariable lower 95% CI" = lower_ci,
                   "Multivariable upper 95% CI" = upper_ci, 
                   "Multivariable p value" = p, 
                   "Univariable effect" = univ_effect, 
                   "Univariable std error" = univ_se, 
                   "Number exposed" = n), 
          "exports/Supplementary Figure 4B data - Antimicrobial class vs Shannon diversity in longitudinal arm.csv", row.names = F)

multivariable_LS_AM_class_diversity_model |> summary()
robust_multivariable_LS_AM_class_diversity_model

# remove temporary variableS
rm(#data_for_LS_AM_class_diversity_model,
   names_of_all_exposures_in_LS_AM_class_diversity_model,
   robust_multivariable_LS_AM_class_diversity_model_data_frame,
   multivariable_LS_AM_class_diversity_model,
   robust_multivariable_LS_AM_class_diversity_model,
   univariable_LS_AM_class_diversity_model_data_frame,
   patient_category_LS_AM_class_diversity_model)
```
