## Supplementary material for "The impact of different antimicrobial exposures on the gut microbiome in the ARMORD observational study": ARMORD R project: SuppTables.R

```
# Data for suplementary tables 1 and 2

## % Abundance of major taxa in baseline sample
c_bugRA |> 
  semi_join(b_first_samples, c("pid", "no")) |> 
  summarise(entc = 100*10^quantile(log_entcRA_trunc, c(0.25,0.5,0.75)),
            entb = 100*10^quantile(log_entbRA_trunc, c(0.25,0.5,0.75)),
            bact = 100*10^quantile(log_bactRA_trunc, c(0.25,0.5,0.75)),
            clos = 100*10^quantile(log_closRA_trunc, c(0.25,0.5,0.75)),
            acti = 100*10^quantile(log_actiRA_trunc, c(0.25,0.5,0.75)))

# Median (IQR) proportion of classified microbiome in the five taxa used for analysis (assigned by Kraken2)
c_bugRA |> 
  mutate(sum_of_major_taxa = 100*(10^log_entcRA_trunc + 10^log_entbRA_trunc + 10^log_bactRA_trunc + 10^log_closRA_trunc + 10^ log_actiRA_trunc)) |> summarise(quantile(sum_of_major_taxa, c(0.25,0.5,0.75)))

# Median (IQR) proportion of classified microbiome in the five taxa used for analysis (assigned by Metaphlan - this was used for diversity metrics but not the individual taxon abundance as limit of detection higher than Kraken2)
Taxa_Metaphlan |> 
  filter(phylum == "Bacteroidetes" | phylum == "Actinobacteria" | class == "Clostridia" | genus == "Enterococcus" | family == "Enterobacteriaceae") |> 
  group_by(samp_id) |> 
  summarise(sum_of_major_taxa = sum(perc)) |> 
  summarise(quantile(sum_of_major_taxa, c(0.25,0.5,0.75)))

## number of samples with major AMR classes detected in any sample
# tetracycline
c_argRA |> filter(log_tet_rpm_trunc > 0.3) |> distinct(pid) |> count()
# beta-lactamase
c_argRA |> filter(log_bla_rpm_trunc > 0.3) |> distinct(pid) |> count()
# aminoglycoside
c_argRA |> filter(log_amg_rpm_trunc > 0.3) |> distinct(pid) |> count()
# macrolide
c_argRA |> filter(log_mac_rpm_trunc > 0.3) |> distinct(pid) |> count()
# vancomycin
c_argRA |> filter(log_van_rpm_trunc > 0.3) |> distinct(pid) |> count()
```
